# Understanding response to treatment in depression: Insights from the Pakistani DIVERGE study

**DOI:** 10.64898/2026.04.13.26350625

**Authors:** Muhammad Umar, Fahad Hussain, Bakht Khizar, Inzemam Khan, Fahadullah Khan, Marius Cotic, Lian Chan, Ashfaq Hussain, Mian Nizam Ali, Shamshad Ahmed Gill, Ali Burhan Mustafa, Imtiaz Ahmed Dogar, Asad Tamizuddin Nizami, Mian Mukhtar ul Haq, Khalid Mufti, Moin Ahmad Ansari, Mian Iftikhar Hussain, Shahzad Tahir Choudhary, Niaz Maqsood, Ghulam Rasool, Hazrat Ali, Muhammad Ilyas, Muhammad Tariq, Sadia Shafiq, Aftab Alam Khan, Saleemuddin Rashid, Hussain Ahmad, Kiramat Ullah Bettani, Muhammad Kamran Khan, Abdul Rashid Choudhary, Muntazir Mehdi, Abdul Shakoor, Nasir Mehmood, Ali Ahsan Mufti, Moti Ram Bhatia, Muhammad Ali, Mir Alam Khan, Naveed Alam, Syed Qalb-i-Hyder Naqvi, Nazish Mughal, Nilofar Ilyas, Parveen Channar, Parveen Ijaz, Allah Din, Huma Agha, Sumaira Channa, Syeda Ambreen, Raza ur Rehman, Gohar Ali, Haider Ali, Sadia Iqbal, Zaheer Ahmad, Sayed Muhammad Sultan, Fahad ul Zain, Dost Muhammad, Zarmina Tareen, Mujeebullah Khan Doutani, Bansi Lal Tanwani, Chooni Lal, Abdul Hadi, Abrar Khan, Moeez Nawaz, Amjad Ali, Asad ur Rehman, Asad Zamir, Basir Muhammad, Cheragh Hussain, Ghulam Haider, Rukhshanda Jabeen, Hina Murtaza Gova, Maira Saleem, Khawaja Hareem Farooq, Khawar Hussain Qureshi, Maqsood Ahmed, Muhammad Abdul Hannan, Muhammad Rehman, Muhammad Saleem, Muhammad Yaqoob Khan, Mujeeb ur Rehman, Naheed Akhtar, Sardar Ali Khan, Shah Fahad, Shaheen Ali, Shoab Shah, Ayesha Javed, Shoukat Ali, Sobia Sabir Ali, Ibrar Ahmed, Ibrar Younas, Jabar Ali, Sohail Tufail, Javed Iqbal, Suresh Kumar, Syed Nazir Ahmad, Wajid Khan, Muhammad Salman Sidiqqui, Muhammad Mohsin Raza, Nick Bass, Andrew McQuiilin, Ayesha Rehman, Syeda Tooba Akbar, Komal Nawaz, Saad Salman, Rida Malik, Kalsoom Nawaz, Sadaf Naureen, Qurrat ul Ain, Sumaira Rajpoot, Dur-e-Nayab, Hina Said, Mahnoor Naeem, Madiha Kanwal, Anbreen Bano, Adan Javed, Samra Khursheed, Aiman Farooq, Ambreen Khan, Anum Saeed, Arshi Rashid, Muhammad Haris Sabir, Isra Waqas, Maida Batool, Qurrat ul Ain, Rabia Ahmad, Sana Fayyaz, Tayyaba Nawaz, Yasmeen Shahwani, Zaib Samraz, Elvira Bramon, Glyn Lewis, Arsalan Hassan, Muhammad Ayub, Karoline Kuchenbaecker

## Abstract

**Background:** Major depressive disorder (MDD), a leading cause of disability worldwide, exhibits substantial heterogeneity in treatment outcomes. Patients who do not respond to standard antidepressant therapy account for the majority of MDD’s disease burden. Risk factors have been implicated in treatment response, including genes impacting on how antidepressants are metabolised. Yet, despite its clinical importance, risk factors for treatment-resistant depression (TRD) remain unexplored in low- and middle-income countries (LMIC). We used data from the DIVERGE study on MDD to investigate the risk factors of TRD in Pakistan.

**Methods:** DIVERGE is a genetic epidemiological study that recruited adult MDD patients (≥18 years) between Sep 27,2021 to Jun 30, 2025, from psychiatric care facilities across Pakistan. Detailed phenotypic information was collected by trained interviewers and blood samples taken. *Infinium Global Diversity Array with Enhanced PGx-8* from Illumina was used for genotyping followed by DRAGEN calling to infer metaboliser phenotypes for Cytochrome P450 (CYP) enzyme genes. We defined TRD as minimal to no improvement after ≥12 weeks of adherent antidepressant therapy. We conducted multi-level logistic regression to test the association of demographic, clinical and pharmacogenetic variables with TRD.

**Findings:** Among 3,677 eligible patients, polypharmacy was rampant; 86% were prescribed another psychotropic drug along with an antidepressant. Psychological therapies were uncommon (6%) while 49% of patients had previously visited to a religious leader/faith healer in relation to their mental health problems. TRD was experienced by 34% (95%CI: 32-36%) patients. The TRD group was characterised by more psychotic symptoms and suicidal behaviour (OR=1.39, 95%CI=1.04-1.84, p=0.02; OR=1.03, 95%CI=1.01-1.05, p=0.005). Social support (OR=0.55, 95%CI=0.44-0.69, p=1.4x10^-7^) and parents being first cousins (OR=0.81, 95%CI=0.69-0.96, p=0.01) were associated with lower odds of TRD. In 1,085 patients with CYP enzyme data, poor (OR=1.85, 95%CI=1.11-3.07, p=0.01) and ultra-rapid (OR=3.11, 95%CI=1.59-6.12, p=0.0009) metabolizers for *CYP2C19* had increased risk of TRD compared with normal metabolisers.

**Interpretation:** There was an excessive use of polypharmacy in the treatment of depression while psychological therapies were uncommon highlighting the need for more evidence-based practice. This first large study of MDD from Pakistan uncovered the importance of culture-specific forms of social support in preventing TRD, highlighting opportunities for interventions in low-income settings. Pharmacogenetic markers can be leveraged to predict TRD.

## Introduction

Major depressive disorder (MDD) is a common psychiatric disorder and a leading cause of disability, with global lifetime prevalence estimates of 13.6% and 7.5% in females and males, respectively.^1–4^ Treatment guidelines recommend antidepressants along with psychological treatment for moderate to severe MDD.^5^ However, there is significant variation in the outcome of an episode after treatment with antidepressant. An estimated 13-38% of patients do not respond to antidepressant treatment, have persistent symptoms and are classified as having treatment resistant depression (TRD).^6,7^ Persistent symptoms reduce productivity and quality of life and can increase feelings of hopelessness in patients.^8–10^ TRD inflicts a substantial economic burden on the society due to higher healthcare resource utilization,^11–13^ along with loss of productivity.^14^ In United States, the estimated national burden of TRD is 47% (43.8 billion USD) of the total diseases burden for MDD.^15^

A widely used criterion in observational studies for TRD is failure of two consecutive adequate trials of antidepressants belonging to different pharmacological classes.^16–18^ Systematic reviews,^19,20^ treatment guidelines,^21^ and consensus statements have adopted this approach. Intervention studies, on the other hand, more commonly counted failure of a single adequate trial as TRD.^22^

Establishing the risk factors for TRD is important to develop measures to reduce risk of TRD and to identify patients at risk of TRD early and adjust their clinical management.^7,23,24^ Early age of onset, comorbid conditions (psychiatric and physical),^6,17,25^ higher suicidal risk, presence of psychotic symptoms^26^ and lower socioeconomic status (SES)^27^ were associated with TRD in previous studies.^28^ However, there is no widely used model to identify patients at risk of TRD.

Genes encoding cytochrome P450 (CYP) enzymes, particularly *CYP2C19* and *CYP2D6*, play a critical role in the metabolism of most prescribed antidepressants. CYP enzyme metaboliser status, which can be inferred from genotypes, has shown corelation with serum drug concentrations,^29–31^ which can lead to differential response to drugs, a key determinant of treatment efficacy and tolerability.^32–34^ The Clinical Pharmacogenetics Implementation Consortium (CPIC) has incorporated this evidence into clinical guidelines, providing *CYP2D6*-and *CYP2C19*-based recommendations for serotonin reuptake inhibitor prescribing.^35,36^

However, despite well-characterized effects on drug metabolism, a consistent association between CYP-mediated metabolism and antidepressant treatment response remains unresolved. Additionally, the frequencies of different metabolizer groups of pharmacogenes vary greatly across populations,^35,36^ underscoring the need for population-specific studies to optimize precision psychiatry approaches.

Most research about TRD has been conducted in Europe and North America and some in East Asia. A recent systematic review investigating the predictors of TRD, synthesized data from 57 studies but did not report any study from South Asia - home to 25 % of world population - nor from any low-middle income countries (LMIC).^37^ Therefore, it remains unclear whether current findings generalise to these regions and whether additional risk factors need to be considered.

Pakistan has a population of 240 million people with unique population characteristics and limited and variable access to psychiatric services.^38^ We aimed to better understand the role of these local cultural, societal and biological factors in treatment response for MDD patients. In this study, we develop a definition of TRD for cross-sectional clinical settings in Pakistan and estimate its frequency. We then investigate risk factors and assess cytochrome P450 (CYP) biomarkers for their association with TRD.

## Methodology

### Study settings and design

The current study used data prom the DIVERGE project (Depression in diverse populations: Unravelling the interplay between genes and environment) which is a multi-site study on the genetic and environmental risk factors for depression in Pakistan.^39^ DIVERGE recruited participants from psychiatric care facilities between 27-09-2021 and 30-05-2025 and, includes 13,393 participants (9,278 MDD cases, 4,115 controls), making it the largest epidemiological study on depression with deep phenotyping and genetics in any non-European population. In Pakistan, the psychiatric care is provided by public and private healthcare setups. Mental health services are not integrated into the primary care setup and are usually available at district headquarters and tertiary care hospitals. Private sector in general includes hospitals and small speciality clinics. The private psychiatric clinics are small psychiatric care facilities managed by individual psychiatrists, which provide out-patient and in-patient services (in some cases). This manuscript is based on data from 8,360 MDD patients that were available when we conducted the analyses. Ethical approval for the DIVERGE study was provided by the National Bioethics Committee Pakistan (ref.no: 4-87/NBC-692/23/1524), University College London Research Ethics Committee (ref.no: 14125/002) as well as local ethics committees at recruitment sites.

### Participants

Detailed patient selection criteria are available elsewhere.^39^ Briefly, MDD cases (≥ 18 years age) from psychiatric outpatient and inpatients departments, in private clinics and public sector hospitals were enrolled into the DIVERGE study via consecutive sampling technique after providing informed consent in writing (Supplementary figure 1). MDD onset above 65 years were excluded. 204 patients refused to participate in the DIVERGE study while 1,902 were excluded because they did not meet inclusion criteria or because they did not demonstrate capacity to participate when approached.

### Procedures

Demographic, psychopathological and treatment related data were collected via a detailed structured interview (Supplementary methods). The interview forms were completed by a trained interviewer, using web-based tool ‘REDCap’, to facilitate participation of illiterate participants.^40^ Treatment response was determined based on how much the patient’s symptoms had changed since starting treatment (Supplementary Table 1). TRD was defined as minimal or no relief in symptoms after 12 or more weeks of use of medication with good/intermediate adherence to medications. Patients who responded to the medications were termed non-resistant. Risk factors reported in peer reviewed journals were selected to test their association with TRD in DIVERGE.^6,7,41^ We also included variables that we considered potentially important with respect to the Pakistani society. These included parental relatedness, available social support as reported by the patient and exposure to traumatic life events (Supplementary Table 2).

### Pharmacogenetic analysis

The study participants were genotyped on the Infinium Global Diversity Array (GDA) with Enhanced PGx-8 v1.0. The microarray data were used to call star alleles for pharmacogenes, using Illumina’s *DRAGEN Array 1.0* package. This array provides 87.6% allele coverage across 19 pharmacogenes which is highest of all available arrays.^42^ The current methodology (GDA+ enhanced PGx) accounts for the copy number variant and tandem arrangements/hybrid alleles of CYP2D6 by incorporating a dedicated PCR. Each call has a likelihood score and an alternative call which can be leveraged as a quality metric for these calls. Individuals were assigned metabolizer phenotype annotations based on diplotypes of the star alleles. We considered two genes, *CYP2D6* and *CYP2C19,* as they metabolize commonly used selective serotonin reuptake inhibitors which are also the most prescribed antidepressants in Pakistani based on our DIVERGE data. The *DRAGEN Array* star allele caller sources the star alleles’ definitions from pharmacogenetic databases including PharmVar and PharmGKB.

### Statistical Analysis

R statistical software v 4.2.1 was used for the data analysis.^43^ Medication compliance was examined and reported for individuals who had a history of psychotropic medication use. To describe the sample, the descriptive statistics such as of mean (SD) and frequency (%) were reported for continuous and categorical variables respectively. A comparative analysis of the of currently prescribed medication data was performed between TRD and NTRD. The frequency of different non-pharmacological and pharmacological therapies along with mean doses were calculated and compared between TRD and NTRD using chi-square/t-test as appropriate. Population frequencies of star alleles and metabolizer phenotypes were estimated for each provincial ethnic group.

To assess the associations of potential risk factors and pharmacogenetic markers, TRD status was used as a binary outcome in logistic regression modelling. Observations from a given clinic might not be fully independent as patients from that site are treated by same clinician and interviewed by same interviewer. To account for this, multilevel logistic regression with random intercept using the *lme4* package in R^44^ was employed to evaluate the associations of the explanatory variables with TRD. Suicidal Behaviour Questionnaire- Revised (SBQ-R) scores were grand mean centred. Univariate multilevel logistic regression was used to assess association of selected variables with TRD status first. To test if the associations from the univariate analyses are robust, we developed separate multivariate models for demographic and clinical variables adjusting for potential confounders and covariates. No multiple testing correction was applied as we relied on the results from multivariate model which is a single test including multiple variables. The same approach was used for testing the association of CYP enzyme metabolizer status with TRD. An association was considered significant if the p-value of the regression coefficient was < 0.05 in the multivariate model. Model performance was assessed using Akaike information criterion (AIC).

To validate the self-reported social support, as measured by the Oslo Social Support Scale (OSSS), we tested the association between OSSS scores and other more objective indicators of social support, such as family structure and relatives living in the same area using ordinal regression and p<0.01 (Bonferroni correction).

## Data availability

The data used for this study are available on a Bonafide research request in line with the study ethics and consent. Data access request can be made at and decision will be taken by the data access committee.

## Code availability

No customized software was used for analysis. Publicly available software and tools cited in the methods were used to conduct data cleaning, statistical analysis, and generate figures. The code will be made available through github.

## Results

### TRD prevalence and sample characterization

Of the 8,360 MDD patients recruited when we carried out the analyses, 1,954 (23%) were medication naïve (MN) and not eligible for this study. The remaining 6,406 reported medication history for depression out of which 41%, 57% and 2% had good, moderate and poor medication adherence, respectively (Supplementary Figure 2). Based on this, 3,677 MDD patients were eligible for inclusion in the TRD analysis (Figure 1). Of these 1,265 (34%) were classified as treatment resistant.

**Figure 1:**
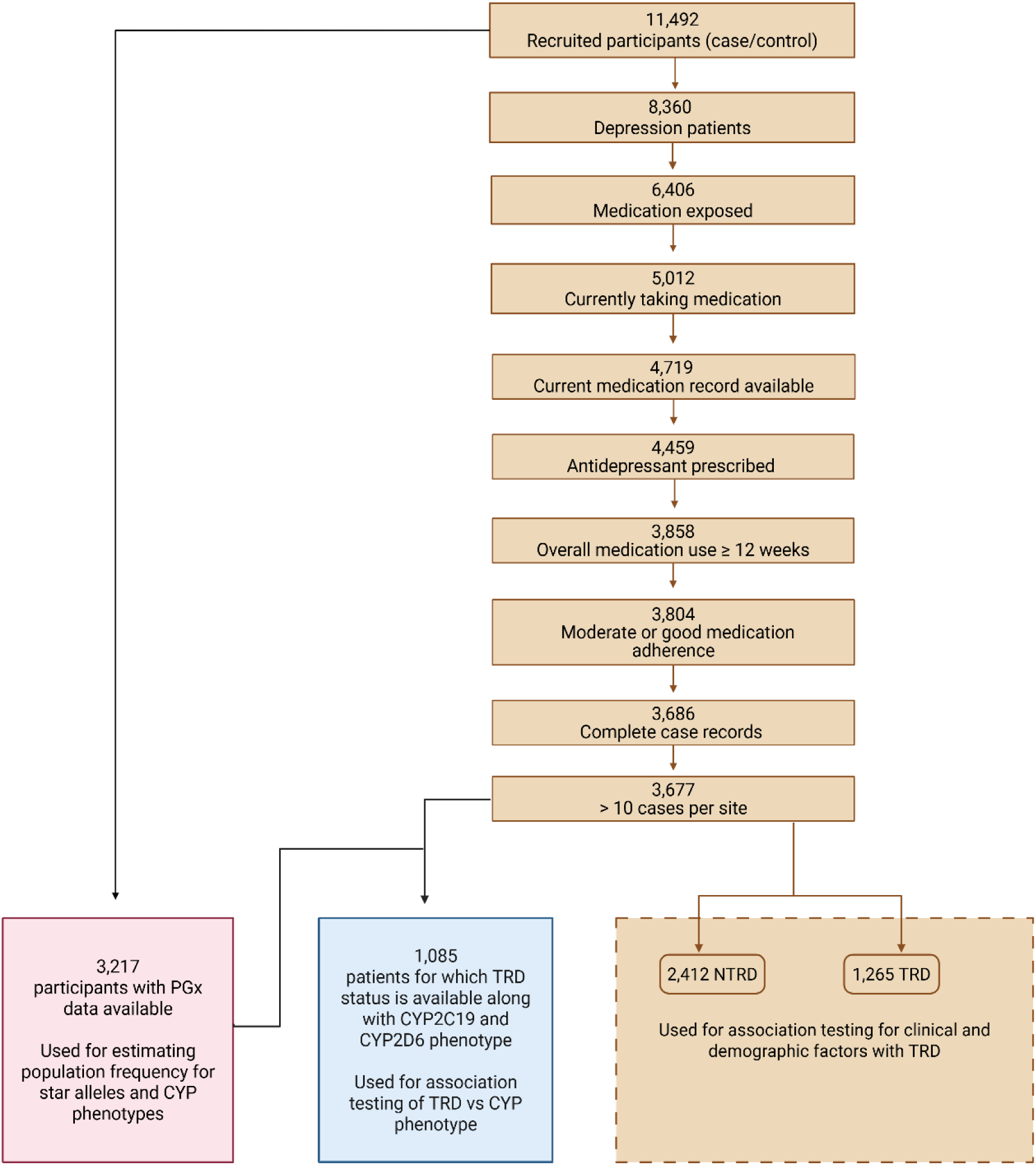
Flowchart showing the study design and patient eligibility criteria formation of the TRD or NTRD groups.

The cohort was predominantly female (56%), with a mean age (Standard deviation=SD) of 38 (11) years and the average duration of illness was 7 years (SD=7). The mean age at onset for depression was 30 years (SD=10). A family history for psychiatric disorders was reported by 39% of participants. Comorbidities were common, including psychiatric (32%) and physical (42%) conditions, notably gastrointestinal (17%) and hypertensive (15%) disorders.

Sociodemographic data revealed high consanguinity rates (45% with parents as 2^nd^ cousin or closer) and significant gender disparities in education with 17% of males vs. 46% of females having never received any formal education. The provincial distribution mirrored Pakistan’s population structure, with 88% of participants from Khyber Pakhtunkhwa or Punjab province (Supplementary Table 3 and 4).

### Psychotropic medications

In the final TRD-eligible medication-exposed patients (N = 3,677), only 14% received a single antidepressant, while the remaining 86% were taking at least one additional psychotropic drug including another antidepressant, a mood stabilizer, antipsychotic, anxiolytic or benzodiazepine (Figure 2, Supplementary Table 5a). In medication naive (MN) patients, this figure was similarly higher with 84% receiving a prescription for a second psychotropic drug. About 29% (MN:21%) of the patients were prescribed combination therapy (> 1 antidepressant) and 52% (MN:39%) augmentation therapy (antidepressant with mood stabilizer or antipsychotic). The use of these drug combinations varied across recruitment sites. Most drug classes and combinations were comparable between the TRD and NTRD groups in terms of frequencies and mean doses (Supplementary Tables 5a and 6).

**Figure 2:**
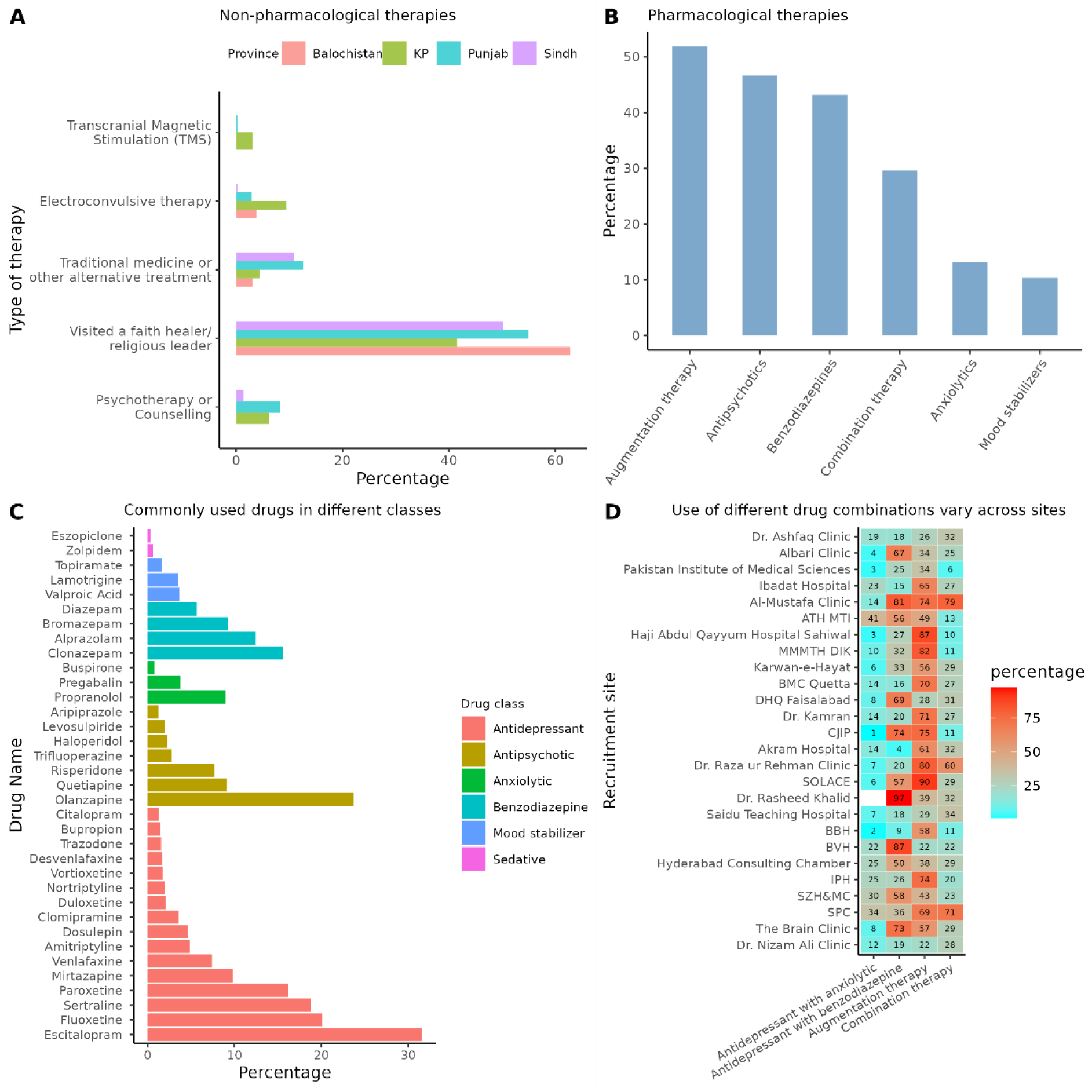
Overview of different therapies and drugs used by the MDD patients (N=3,677), reported by provincial ethnicity and recruitment site. A) Bar plot comparing proportions of non-pharmacological therapies reported by the DIVERGE MDD patients, shown separately for different provinces distinguished by colour. B) The frequency of use of different pharmacological therapies . Augmentation therapy = antidepressant along with antipsychotic or mood stabilizer. Combination therapy = more than 1 antidepressant. C) Current drugs by pharmacological drug-class (colour-coded). D) Heatmap depicting the frequency of use of different drug-class combinations across recruitment sites. X-axis showing drug combinations and y-axis has recruitment sites. The sites are arranged in the increasing order of log odds of TRD from multilevel logistic model with no predictors (null multilevel logistic model with random effects added at site level and no predictors included).

### Non-pharmacological management

The most common among the non-pharmacological strategies was visiting a religious leader/faith healer (49%). The use of traditional/alternative medicine was reported by 6% of the patients. The use of psychotherapy and electroconvulsive therapy was about 6% each. Psychotherapy was reported significantly more frequently by the NTRD (7%) compared to the TRD (4%) group (p=0.0003) (Supplementary Table 5a).

### Risk factors for TRD

Multilevel logistic regression with random intercept demonstrated better fit to the data than logistic regression (p<2.2×10^-16^),with ‘site’ explaining 8% of variance in the TRD measured by interclass correlation coefficient (Supplementary Figure 3, Supplementary Table 7).

A significant association was observed between social support and TRD where patients with moderate and strong social support showed reduced odds of TRD by 36% and 45% respectively (OR=0.64, 95% CI=0.53-0.76, p=9.8 x 10^-7^ ; OR=0.55, 95% CI=0.44-0.69, p=1.4 x 10^-7^). To validate this finding, we assessed whether people reporting higher social support on OSSS have comparatively better support structures available to them (Supplementary Table 8). Results show that moderate or high social support was more frequently reported by patients who had other relatives living in the same area compared to no relatives (OR=4.20, 95% CI=3.60-4.95, p=3.9 x 10^-71^). Additionally, a larger family structure (joint or nuclear) with higher number of individuals in the family household was also linked to higher perceived social support (Supplementary Table 8).

Parents being first cousin was more common in the NTRD group (39% vs34 %) and was associated with a reduced odds of TRD by 19% (OR=0.81, 95% CI=0.69-0.96, p=0.01). Divorce was more frequent in TRD than NTRD (3.5% vs 2.5%, OR=1.81, 95% CI =1.18-2.78, p=0.007).

### Differences in clinical characteristics between responders and non-responders

In terms of clinical presentation psychotic symptoms and suicidal behaviour score were higher in the TRD group compared with the NTRD group (OR=1.39, 95% CI=1.04-1.84, p=0.02; OR=1.03, 95% CI=1.01-1.05, p=0.005), while, family history for common psychiatric disorders was associated reduced odds of TRD (OR=0.84, 95% CI=0.71-0.98, p=0.037).

The multivariate model including all variables explained 14% of variance on the liability scale. However, in this full model, inclusion of marital status, parental relatedness and provincial ethnicity did not significantly improve the model fit (Supplementary Table 9a-9b, Supplementary Figure 4).

### Association of pharmacogenetic markers with TRD

Star alleles and metabolizer phenotypes for *CYP2D6* and *CYP2C19* were available for 3,217 participants in DIVERGE. We used these data to report the star alleles and metabolizer phenotypes frequencies in Pakistani population and compared these against other ancestral populations available from PharmGKB (Supplementary Table 10-12). The frequency of *CYP2C19* poor metabolizers in Pakistan was 8% (East Asian 13%, European 2%) (Supplementary Figure 5-6). In Pakistan, there were notable interprovincial differences (Figure 3). For *CYP2C19*, ultrarapid metabolizer were almost 4 times more abundant in Khyber Pakhtunkhwa than Sindh (5.6% vs 1.5%), however, the poor metabolizer frequency in Khyber Pakhtunkhwa (5.6%) was half of that in Punjab (10.5%).

**Figure 3:**
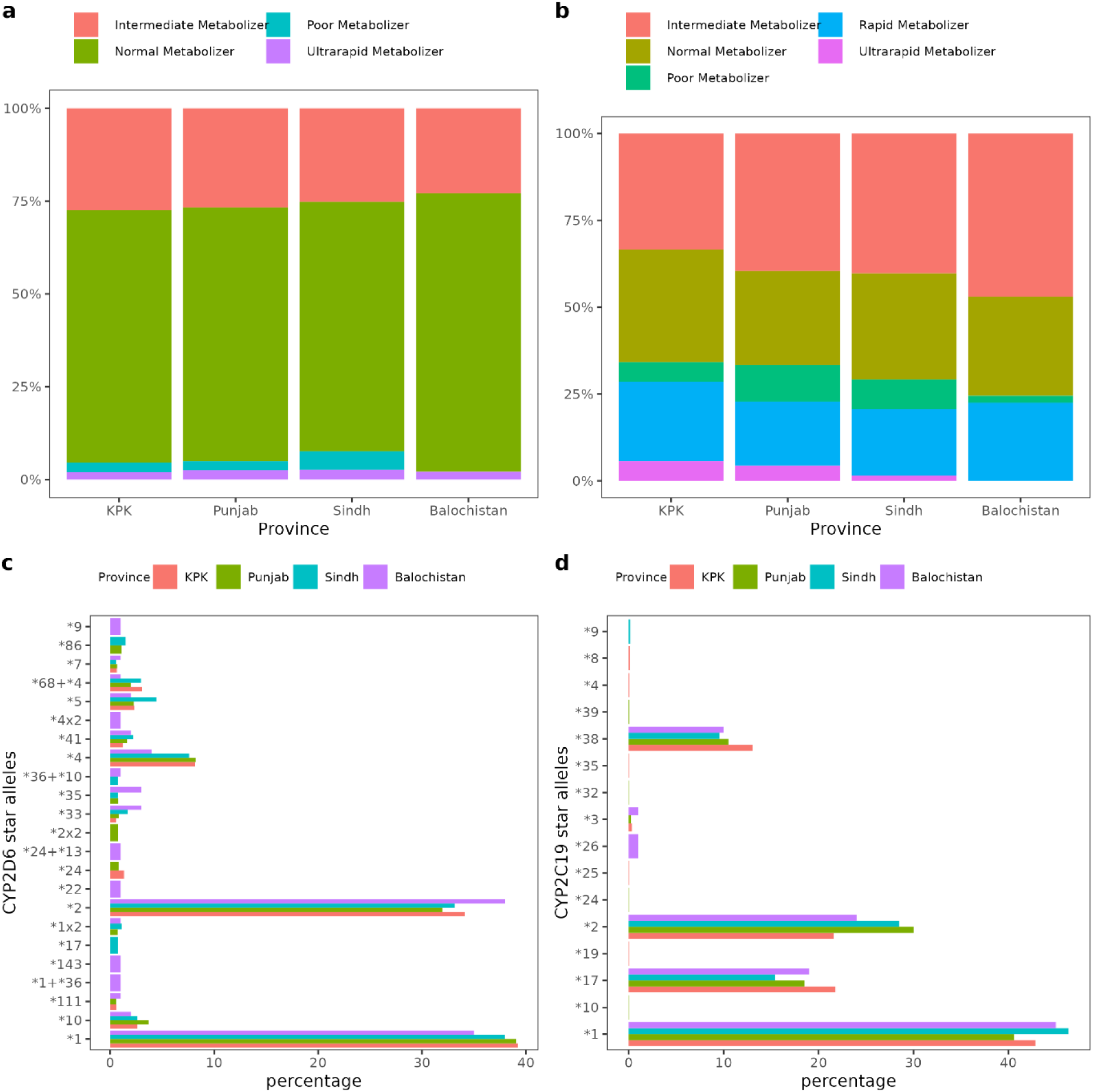
Frequency of star alleles and metabolizer status for *CYP2C19* and *CYP2D6* by province. Plot showing the frequencies of metabolizer phenotypes and star alleles across 3,217 individuals. from different Pakistani provinces a) metabolizer phenotype distribution for *CYP2D6*, b) metabolizer phenotype distribution for *CYP2C19*, c) star allele distribution for *CYP2D6*, d) star allele distribution for *CYP2C19*.

For *CYP2D6* poor metabolizers were most prevalent in Sindh (4.8 %) and least prevalent in Khyber Pakhtunkhwa (2.4%) (Supplementary Table 13-14). Out of the 3,217 participants with metabolizer status, 1,085 overlapped with those eligible for the TRD analysis. Ultra-rapid (OR=3.11, 95% CI=1.59-6.12, p=0.0009), intermediate (OR=1.41, 95%CI=1.00–1.97, p=0.048) and poor (OR=1.85, 95% CI=1.11-3.07, p=0.01) metabolisers for *CYP2C19* were associated with higher odds of TRD compared to normal metabolisers (Table 1). We found no evidence for a *CYP2D6* metabolizer status effect on TRD likelihood. We explored the link between reported side-effect and metabolizer status of *CYP2C19* and *CYP2D6* but found no evidence of association (Supplementary Table 15).

**Table 1:**
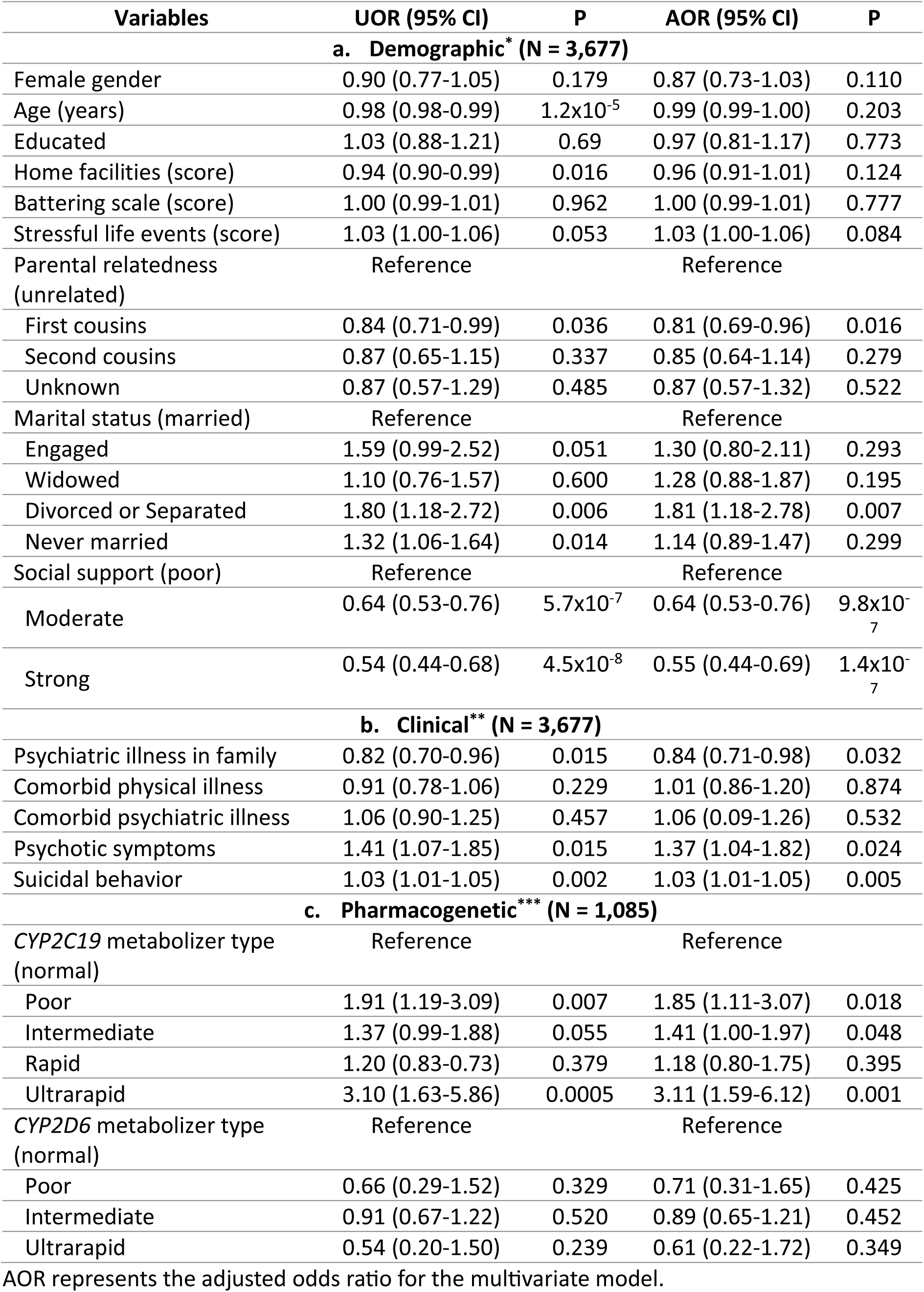
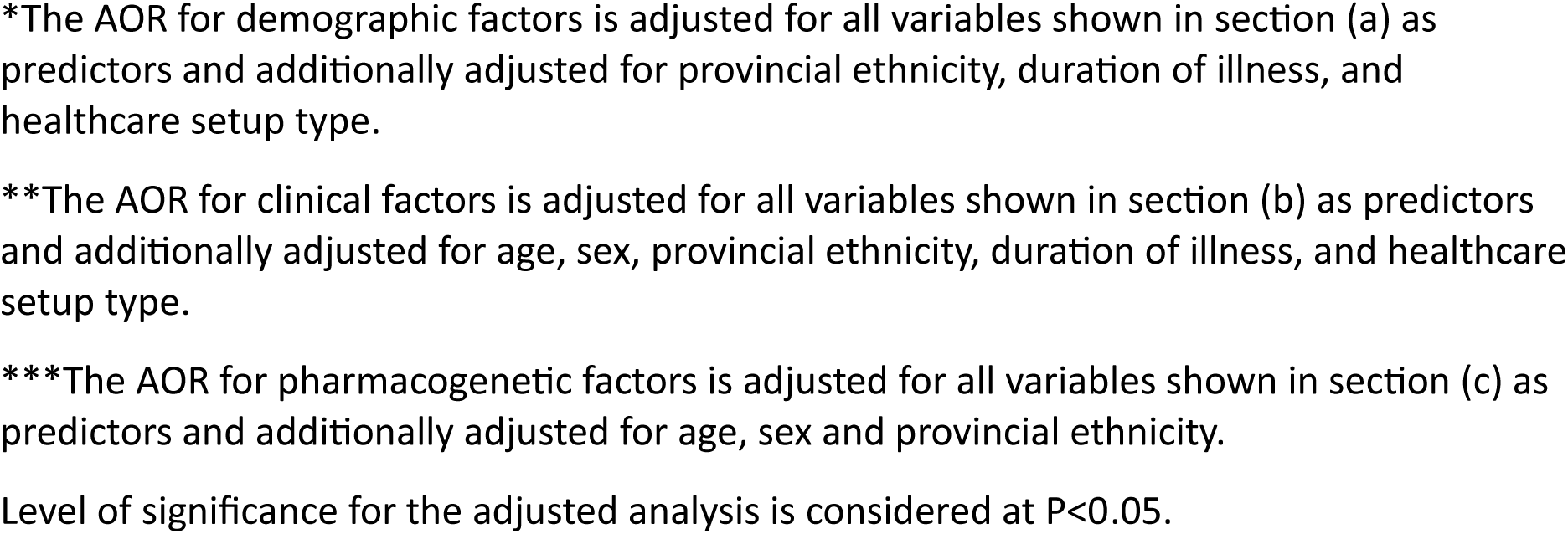
Factors associated with TRD.

| Variables | UOR (95% CI) | P | AOR (95% CI) | P |
| --- | --- | --- | --- | --- |
| **a. Demographic\* (N = 3,677)** |  |  |  |  |
| Female gender | 0.90 (0.77-1.05) | 0.179 | 0.87 (0.73-1.03) | 0.110 |
| Age (years) | 0.98 (0.98-0.99) | 1.2x10<sup>-5</sup> | 0.99 (0.99-1.00) | 0.203 |
| Educated | 1.03 (0.88-1.21) | 0.69 | 0.97 (0.81-1.17) | 0.773 |
| Home facilities (score) | 0.94 (0.90-0.99) | 0.016 | 0.96 (0.91-1.01) | 0.124 |
| Battering scale (score) | 1.00 (0.99-1.01) | 0.962 | 1.00 (0.99-1.01) | 0.777 |
| Stressful life events (score) | 1.03 (1.00-1.06) | 0.053 | 1.03 (1.00-1.06) | 0.084 |
| Parental relatedness (unrelated) | Reference |  | Reference |  |
| First cousins | 0.84 (0.71-0.99) | 0.036 | 0.81 (0.69-0.96) | 0.016 |
| Second cousins | 0.87 (0.65-1.15) | 0.337 | 0.85 (0.64-1.14) | 0.279 |
| Unknown | 0.87 (0.57-1.29) | 0.485 | 0.87 (0.57-1.32) | 0.522 |
| Marital status (married) | Reference |  | Reference |  |
| Engaged | 1.59 (0.99-2.52) | 0.051 | 1.30 (0.80-2.11) | 0.293 |
| Widowed | 1.10 (0.76-1.57) | 0.600 | 1.28 (0.88-1.87) | 0.195 |
| Divorced or Separated | 1.80 (1.18-2.72) | 0.006 | 1.81 (1.18-2.78) | 0.007 |
| Never married | 1.32 (1.06-1.64) | 0.014 | 1.14 (0.89-1.47) | 0.299 |
| Social support (poor) | Reference |  | Reference |  |
| Moderate | 0.64 (0.53-0.76) | 5.7x10<sup>-7</sup> | 0.64 (0.53-0.76) | 9.8x10<sup>-7</sup> |
| Strong | 0.54 (0.44-0.68) | 4.5x10<sup>-8</sup> | 0.55 (0.44-0.69) | 1.4x10<sup>-7</sup> |
| **b. Clinical\*\* (N = 3,677)** |  |  |  |  |
| Psychiatric illness in family | 0.82 (0.70-0.96) | 0.015 | 0.84 (0.71-0.98) | 0.032 |
| Comorbid physical illness | 0.91 (0.78-1.06) | 0.229 | 1.01 (0.86-1.20) | 0.874 |
| Comorbid psychiatric illness | 1.06 (0.90-1.25) | 0.457 | 1.06 (0.90-1.26) | 0.532 |
| Psychotic symptoms | 1.41 (1.07-1.85) | 0.015 | 1.37 (1.04-1.82) | 0.024 |
| Suicidal behavior | 1.03 (1.01-1.05) | 0.002 | 1.03 (1.01-1.05) | 0.005 |
| **c. Pharmacogenetic\*\*\* (N = 1,085)** |  |  |  |  |
| CYP2C19 metabolizer type (normal) | Reference |  | Reference |  |
| Poor | 1.91 (1.19-3.09) | 0.007 | 1.85 (1.11-3.07) | 0.018 |
| Intermediate | 1.37 (0.99-1.88) | 0.055 | 1.41 (1.00-1.97) | 0.048 |
| Rapid | 1.20 (0.83-0.73) | 0.379 | 1.18 (0.80-1.75) | 0.395 |
| Ultrarapid | 3.10 (1.63-5.86) | 0.0005 | 3.11 (1.59-6.12) | 0.001 |
| CYP2D6 metabolizer type (normal) | Reference |  | Reference |  |
| Poor | 0.66 (0.29-1.52) | 0.329 | 0.71 (0.31-1.65) | 0.425 |
| Intermediate | 0.91 (0.67-1.22) | 0.520 | 0.89 (0.65-1.21) | 0.452 |
| Ultrarapid | 0.54 (0.20-1.50) | 0.239 | 0.61 (0.22-1.72) | 0.349 |

AOR represents the adjusted odds ratio for the multivariate model.
\*The AOR for demographic factors is adjusted for all variables shown in section (a) as predictors and additionally adjusted for provincial ethnicity, duration of illness, and healthcare setup type.
\*\*The AOR for clinical factors is adjusted for all variables shown in section (b) as predictors and additionally adjusted for age, sex, provincial ethnicity, duration of illness, and healthcare setup type.
\*\*\*The AOR for pharmacogenetic factors is adjusted for all variables shown in section (c) as predictors and additionally adjusted for age, sex and provincial ethnicity.
Level of significance for the adjusted analysis is considered at $P < 0.05$ .

## Discussion

With 3,677 treatment-exposed MDD patients from across Pakistan, this is the first large observational study of TRD in a clinical setting from an LMIC, to the best of our knowledge.

Amongst MDD patients on antidepressants presenting to psychiatric care facilities in Pakistan, one third did not respond to treatment. We identified stark differences in prescribing patterns across recruitment sites and demonstrated the overreliance on pharmacological compared with psychological therapies for treatment of depression in this setting. Patients experiencing TRD were characterised by higher suicidal ideation and more psychotic symptoms. *CYP2C19* metabolizer status and lack of social support were risk factors for TRD.

Amongst the medication-exposed patients encountered in these clinics and hospitals across Pakistan 34% (95% CI=32%-36%) were treatment resistant. However, the retrospective design may impact these estimates, for example through ascertainment bias. Similar rates were previously reported by other studies in Europe and the USA that employed longitudinal study designs.^6,25^ It has been observed that studies based on administrative databases such as electronic health records and prescription claims databases generally report lower estimates for TRD.^7,23,24,27^ This variation is attributable to differences in TRD definition, data source and population specific factors among studies. About 8% of the variation in TRD liability was attributed to site effects in this study. Differences in prevalence rates of TRD could be linked to local differences in the distributions of risk factors or in prescribing patterns of clinicians leading to different response rates.

The high use of concomitant psychotropic drugs alongside ADs made the implementation of two trial failure definition for TRD impractical in this retrospective cross-sectional study, as only 14% received a single AD alone. For example, the use of antipsychotics as an adjunct in our sample was much higher (47%) in comparison to depression patients in UK primary care (3%) and US based studies (14-21%).^45–47^ This high rate of concomitant medications with AD was also present in medication naïve patients where 50% of the patients were prescribed another psychotropic drug (antidepressant/antipsychotic/mood stabilizer) along with an AD. The prescribing practices with respect to these medications varied significantly across recruitment sites. Possible reasons for the high use of concomitant psychotropic drugs along with antidepressant could include 1) a more severe clinical form of depression, since people with mild depression are unlikely to seek treatment 2) the local prescribing culture which could be influenced by many other factors, 3) a high rate of treatment failure. This high percentage of polypharmacy highlights an urgent need for further research in these clinical settings in Pakistan to understand the factors leading to these prescribing patterns which can inform policies to support evidence-based treatment strategies in this population. Promoting rational prescribing also bears pharmacoeconomic benefits.

Provision of evidence-based psychiatric care can face challenges related to patients’ beliefs about illness, financial constraints, lack of resources and infrastructure, scarcity of trained professional staff, lack of policy guidelines and consequent implementation, and public awareness.^48^ Nearly half of the DIVERGE participants reported a visit to a faith healer or religious cleric over the course of their illness, which is generally a common practice in many Muslim communities.^49,50^ Despite the evidence of being beneficial in depression, only 6% of the patient population in total received some form of psychotherapy which is significantly less than that in western countries (23-43%).^51–53^

One theme emerging from our findings is the association of indicators of social support with treatment outcome. For example, the strongest risk factor for TRD was social support measured by OSSS, where high social support reduced odds of TRD by 45%. The strong correlation of self-reported social support with more objective measures, such as family proximity, makes it unlikely that the link with TRD is explained entirely by negative views of social support rather than actual lack of support. Prior evidence from the literature supports our finding where pre-treatment higher social support was linked to remission or better treatment outcomes in depression.^54,55^

Divorce is uncommon in Pakistan (<1%) and it can lead to increased stress and decreased social support.^56^ 3.5% of patients with TRD reported divorce or separation in comparison to 2.5% in the NTRD group. Previous research has shown that individuals who were divorced had more severe and recurrent form of depression.^57–59^

Finally, the protective effect of parents being first cousins with respect to TRD could also be explained by more support available in such family structures. The couple in a consanguineous union is often surrounded by relatives and elders and it has been reported that this might offer a sense of support and better conflict management.^60^ However, there is need to understand the possible mechanism involved in social support pathways in the context of Pakistani society.

We found the clinically distinctive features between TRD and NTRD where psychotic features and suicidal behaviour were associated with TRD. These results are consistent with previous research and can potentially be used as a predictor for early identification and subsequent management of TRD patients.^13,18,61^

The commonly used SSRIs and tri-cyclic antidepressants are metabolized by *CYP2C19* and *CYP2D6* enzymes. The frequency of these CYP enzymes vary across ancestral populations. Knowing these frequency differences can help identify populations at higher risk of differential response to certain drugs. *CYP2C19* and *CYP2D6* not only metabolize antidepressants but also other important drugs used in the management of psychiatric disorders, pain, cardiovascular disorders and gastrointestinal disorders. This study provides a useful, resource for others, showing the distribution of star alleles and metaboliser phenotypes for *CYP2C19* and *CYP2D6* across Pakistan in a large sample. Frequencies in DIVERGE were similar to those of South/Central Asians but different from Europeans and East Asians. Moreover, we found a significant difference in the frequencies of metabolizer phenotypes of these genes across provincial groups showing a north to south gradient across the country. This was particularly evident for the poor metabolizers in both genes as Sindh had considerably higher frequencies of these compared to other provincial groups.

Whilst it is a plausible hypothesis that metabolizer status influences response to antidepressant medication, other large studies failed to identify an association between *CYP2C19* and treatment response in MDD.^62–65^ We found that ultrarapid and poor metabolizers for *CYP2C19* had a significantly increased risk of TRD The association between being an ultrarapid metabolizer with increased risk of TRD is aligned with the notion that these individuals have decreased drug exposure and thus higher risk of treatment failure.^30,31^ A possible explanation for the *CYP2C19* poor and intermediate metabolizer status leading to increased odds of TRD could be the higher rate of adverse drug reactions which can lead to discontinuation or frequent switching and shorter duration of treatment for a specific drug.^66^ We tested this only for the current medications but found no link between *CYP2C19* and reported adverse reactions. However, data on side effects were not available over the entire course of the illness, and it is possible that the frequent use of concomitant psychotropic medications influenced the reporting of side effects.

Studying TRD in a cross-sectional setting limits our ability to establish temporal relationships between factors that are tested for their association with TRD. DIVERGE is a genetic epidemiological study in an LMIC setting where the cross-sectional approach is most cost-effective and feasible. Data were collected retrospectively through interview; this could potentially introduce a recall bias. We employed the use of multilevel logistic regression to control for site effects. Missingness of prescription records for 300 patients led to the exclusion of subjects from the study. Conducting an analysis to assess the effects of *CYP2C19* and *CYP2D6* on individual drugs was not possible as TRD was defined over the course of the illness and only current prescription data were sufficiently complete. Other non-psychotropic medications can inhibit the activity of *CYP2C19* and *CYP2D6* and thus alter its phenotype, a process known as phenoconversion. No data were available for non-psychiatric medications to account for this. Lastly, we did not follow the two-trial failure definition of TRD, as implementing that in our study was not possible.

Our study uncovered the immense burden of TRD in Pakistan. There are stark differences in the pharmacological management of MDD in comparison to high income countries. We show that risk factors for TRD are shaped by the local cultural context and research in diverse populations can be helpful in identifying culture specific interventions aimed at precision psychiatry in depression. There was evidence that strong support structures are the most important factor associated with treatment outcomes for MDD in Pakistan. Pharmacogenetic markers can be leveraged to predict TRD. This study identified key areas for further research and can serve as a guide for future observational or interventional research in this area.

## Data Availability

All data produced in the present study are available upon reasonable request to the author

