## Supplementary material for "Understanding response to treatment in depression: Insights from the Pakistani DIVERGE study"

**Supplementary Information**

### 1. Supplementary Methods

#### 1.1 DIVERGE study

DIVERGE is a case control genetic epidemiological study of depression in Pakistan that concluded recruitment on 30th June 2025. The study description and protocol in detail is available elsewhere but we provide brief background here for the readers [1]. Starting on 27th September 2021, DIVERGE has undertook recruitment in more than 61 sites across Pakistan. These sites spanned over 4 major provinces of Pakistan and covered all the major ethnic groups in the country. The participants were asked about their demographics, socioeconomic status, disease symptomatology, psychiatric treatment history, psychiatric family history, treatment response and side effects, suicidal behaviour, stressful life events and experiences with battering. Social support available to them was also assessed. Some of the sections were specific to the female sex with questions related to menstrual health, pregnancies, and depression in pregnancy.

##### Inclusion and exclusion criteria for MDD cases

The main inclusion criterion for cases was a lifetime MDD diagnosis according to The Diagnostic and Statistical Manual of Mental Disorders - fifth edition (DSM-V) or DSM-IV or International Classification of Diseases, tenth revision (ICD-10) and being adult (age = 18 years or older) [2-4]. Any prior neurological condition or head injury in close temporal proximity to the MDD onset as well as psychiatric conditions like schizophrenia, bipolar affective disorders, psychotic symptoms preceding depression onset and MDD onset after 65 years of age are exclusion criteria.

##### Setting

Cases were enrolled mostly from psychiatric outpatient departments (OPDs) and inpatients at few sites, in private clinics and public sector hospitals. These enrollment sites are present across the country. The electronic health record keeping in Pakistan is limited to private hospitals mostly and some public sector hospitals and is even rarer in private clinics. There is no integration of these records into one single system across different healthcare setups which could be used for retrieving data for the purpose of research. Patients visiting psychiatrists would have to carry their prescription record along with any psychiatric history notes with them on hard paper files. In DIVERGE the data were collected retrospectively via a detailed interview with the patient. The medication data were obtained by going through the prescription record available with the patient and recording it onto the database used for data collection. This makes it difficult to obtain complete data on medications as not everyone has this record on them while visiting the doctor and, was one of the reasons for excluding individuals from the analysis of treatment response.

#### 1.2 Procedures

##### Enrollment process

Patients are enrolled on their routine visits to the doctor. After, an initial invitation from clinician to participate in the study is accepted by the potential participant, they get referred to the interviewer -a clinical psychologist who had received training about the interview questionnaires and procedures. Participants were further informed about the aims of the study and the potential risks involved and that their participation is completely voluntary. The interview began only after the participant was consented in writing. The schematic of the enrollment process is available ( **Supplementary** figure **1)**.

##### Patient assessment

For broad psychopathological assessment and confirmation of the depression diagnosis, a mental health screener alongside the Diagnostic interview for Psychosis and Affective Disorder (DI-PAD) were used. The mental health screener and DI-PAD were initially developed for genetic epidemiological studies, primarily in the US, for the Genomic Psychiatry Cohort (GPC) [5] and later adapted for another genetic study in Pakistan [6]. The screener collects information on mental health, physical health, consanguinity, demographics, and ethnicity, while the DI-PAD is used to confirm the MDD diagnosis and collect data on psychiatric family history and age of onset.

The Oslo Social Support-3 (OSSS-3) was used to measure social support. It generates scores ranging from 3-14. Based on these scores 3 social support categories were formed, i.e poor (3-8), moderate (9-11), strong (12-14). Life Events Checklist-5 (LEC-5) [7] was used for recording stressful life events. Each item in the LEC-5 has multiple options which are scored as shown here (Happened to me = 3, witnessed it =2, happened to close family/friend = 1, part of job =

2, not sure/does not apply = 0). The LEC-5 score for overall exposure is generated by summing up individual item score to give a single score. Women Experiences with Battering (WEB) scale was adapted to be usable for all participants independent of gender[8]. Suicidal Behaviour Questionnaire-R (SBQ-R) was used for screening suicidal behaviour. This was adapted for the DIVERGE study. The adapted form would generate scores from 1-12 as the increasing score would indicate high severity in suicidal behaviour[9]. During the interview patients were asked about family history -up to 3rd degree relatives- of common psychiatric disorders. These disorders included depression, schizophrenia, bipolar disorder, obsessive compulsive disorder and anxiety disorder. Based on this information, a new variable called 'psychiatric family history' was derived which recorded the presence of any self-reported positive psychiatric family history or total absence of psychiatric family history at all. DIVERGE collected information on different psychiatric and physical health conditions. However instead of separately testing for all of them for association with treatment resistant depression (TRD), we derived one binary variable (yes/no) each for psychiatric or physical comorbidities as a group, that marked the presence/absence of any psychiatric or physical comorbidity respectively. The educational status which is binary was derived from a categorical variable which records the level of education, if obtained, from primary up to post-graduate. Home facilities score was generated as an indicator for the socioeconomic status. It is based on the absence or presence of certain facilities in the home. The presence was scored as '1' and absence '0'. All eight items were then summed up to give a final score that was used as a raw indicator of the socioeconomic status of participants in the study.

#### 1.3 Recording medication information and response to medication

In the treatment related portion of the interview forms, DIVERGE study recorded the current, past and new prescriptions, if available. These were essentially prescriptions different from each other. Interview forms have a dedicated section for recording response to medications called 'Treatment Response History'. It asked questions on overall change in symptoms since start of treatment, duration of treatment received and medication compliance.

Response to treatment in DIVERGE was assessed based on the patient's rating of improvement in symptoms since the start of treatment. Those who responded saying they experienced minimal or no relief at all were termed as resistant, while the rest were considered non-resistant.

#### 1.5 Statistical analysis

##### Rationale for fitting random intercept but not random slope

Random intercept and fixed slope multilevel logistic regression model was fit to the data. Since, investigating the overall effect estimates (odds ratios) for the predictor variables were desired for this analysis, so allowing slopes to vary across sites did not seem rational. However, as sensitivity analysis, random slope models were fit for every predictor variable one at a time and compared to random intercept-fixed slope model via likelihood ratio tests, but it did not improve the model.

##### Multilevel logistic regression

The prescribing practices among the clinicians at different sites varied considerably. Additionally, the rating of the response to medication item could be affected by interviewer bias, this could potentially affect the outcome on part of the patient. So it is hypothesised that the odds of a patient to be resistant would vary across clinics, hence we fit random intercept. As we were interested in the overall effect of the explanatory variables, we did not allow the slope to vary across sites. Level one was the 'patients' while level two was the hospitals and clinic i.e., 'Site' from where these patients were enrolled into the study.

Before testing variables for association multicollinearity was checked using variance inflation factor (VIF) and variables with values above 5 were not included in the multivariate model. Due to collinearity with other variables, 'onset age' and 'clinic provincial location' were excluded in the final model. Null models for simple logistic regression as well as multilevel logistic regression were fitted, and ANOVA was run for model comparison to test model fit to the data.

#### 2. Supplementary Results:

##### 2.1 Individual Drug Report

The comparison of frequency of individual drugs and their mean doses for resistant and the non-resistant group is given (Supplementary Table 6). For most of the drugs used, the two groups do not differ significantly in frequencies or mean doses except a few. Fluoxetine, Dosulepin and Lorazepam were more frequently found in the non-treatment resistant depression (NTRD) group but there was no significant difference in the mean dose between the groups for these drugs. Desvenlafaxine, Trazodone, and Zolpidem were significantly higher in frequency in the TRD vs NTRD with no significant difference in mean doses. Additionally, the mean doses of Sertraline and Clonazepam were higher in the NTRD group than the TRD group. We did not correct for multiple comparison as the main aim behind this was to show that the medications received by both the groups are comparable.

#### 3. Supplementary Figures

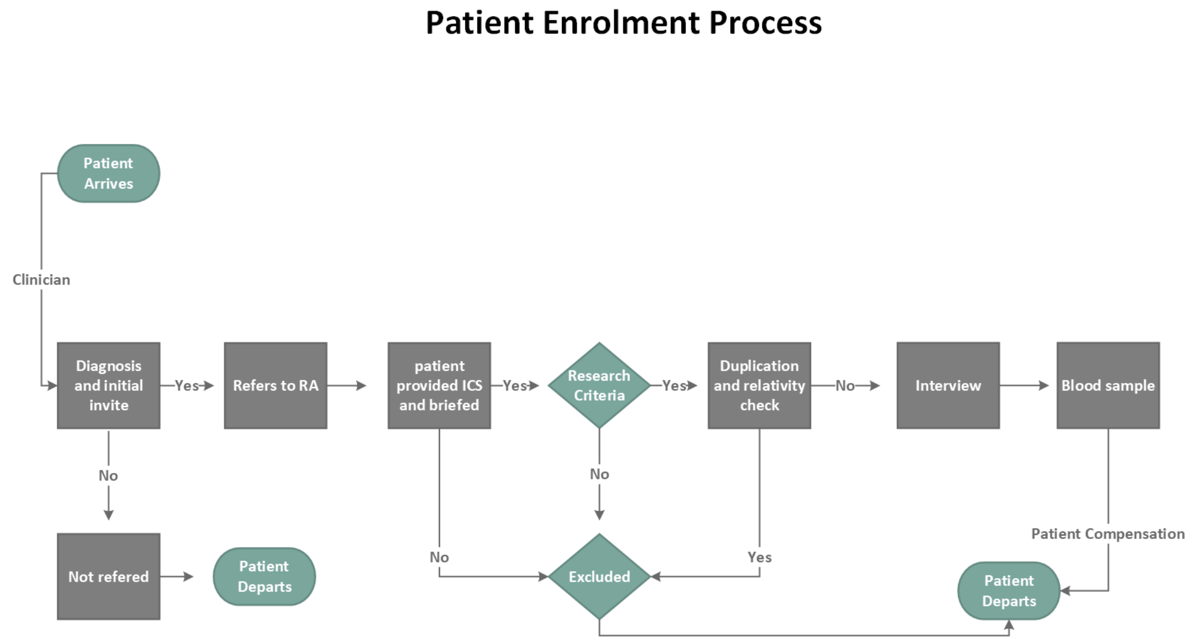

**Supplementary Figure 1:** Flowchart of patient enrolment process at outpatient clinics

Abbreviations: RA = research assistant, ICS = informed consent sheet.

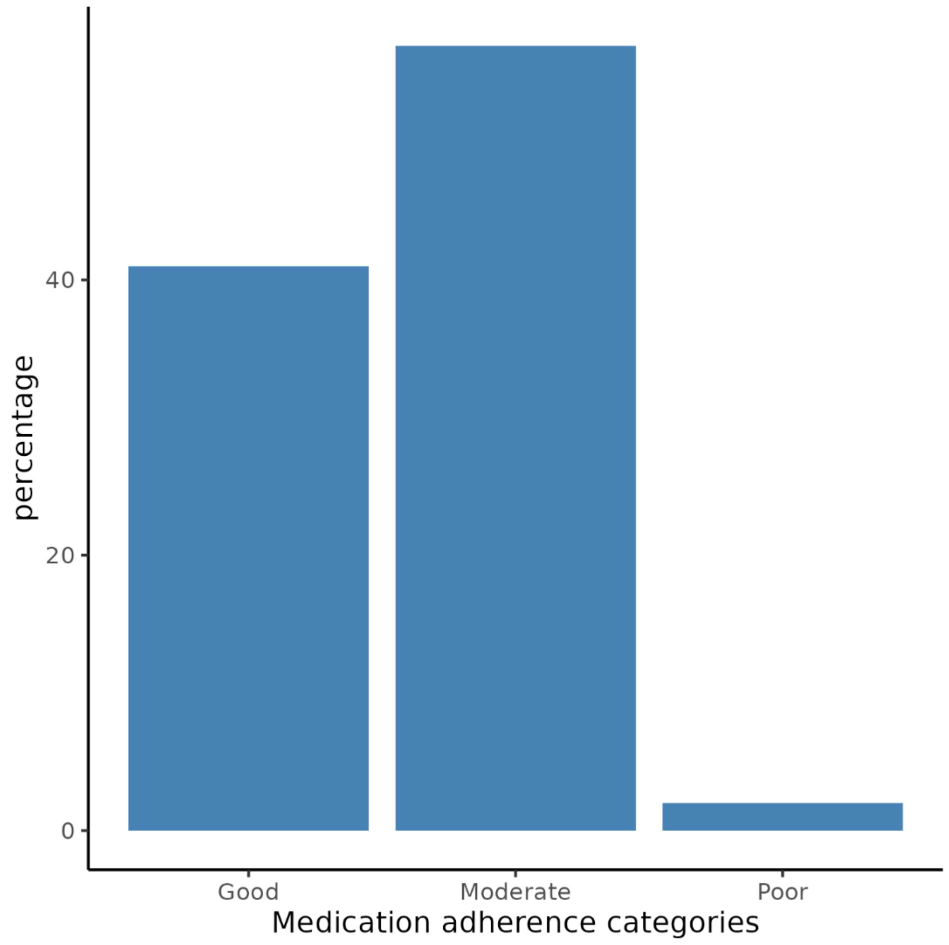

**Supplementary Figure 2:** Medication adherence in MDD patients with medication history. The adherence to medication is reported in individuals who have psychiatric medication history for depression in DIVERGE (N = 6,406). Medication adherence was recorded via a single question which allows to select multiple options. It generates scores from 0-4, which were grouped into good (0), moderate (1-2) and poor (> 2) categories.

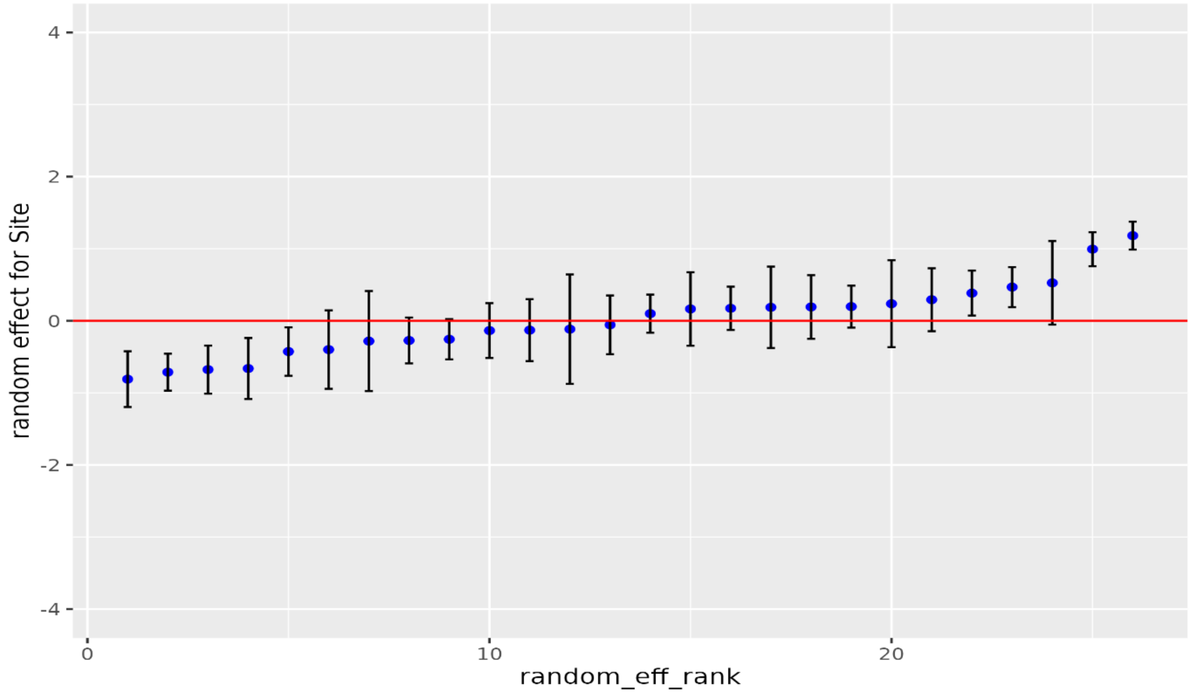

**Supplementary Figure 3:** Plot represents the random effects variation among sites. X-axis show random effect rank which is a number given to the sites in sequence after arranging them in order of increasing log odds of TRD from null multilevel logistic regression model. The Y-axis represents the random effects in terms of z-scores. The blue dot represents z-score converted log odds from null multilevel model for a specific site while bars around it show standard error.

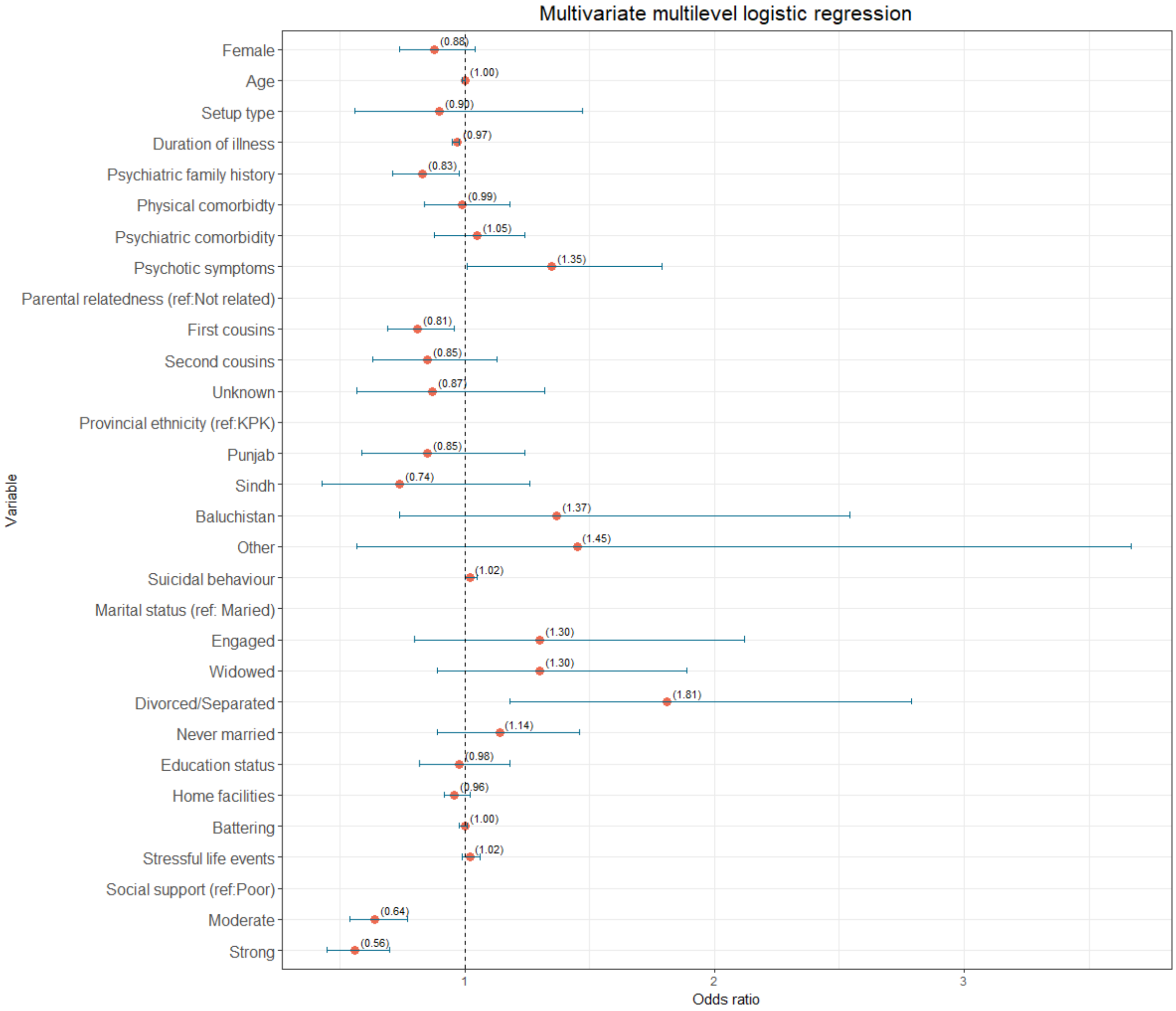

**Supplementary Figure 4:** TRD risk factors. Forest plot depicts results of the full regression model (N=3677). The dot represents the adjusted odds ratio while the bars on both sides of the dot show 95 % confidence intervals. The doted vertical line represents the odds ratio of 1.

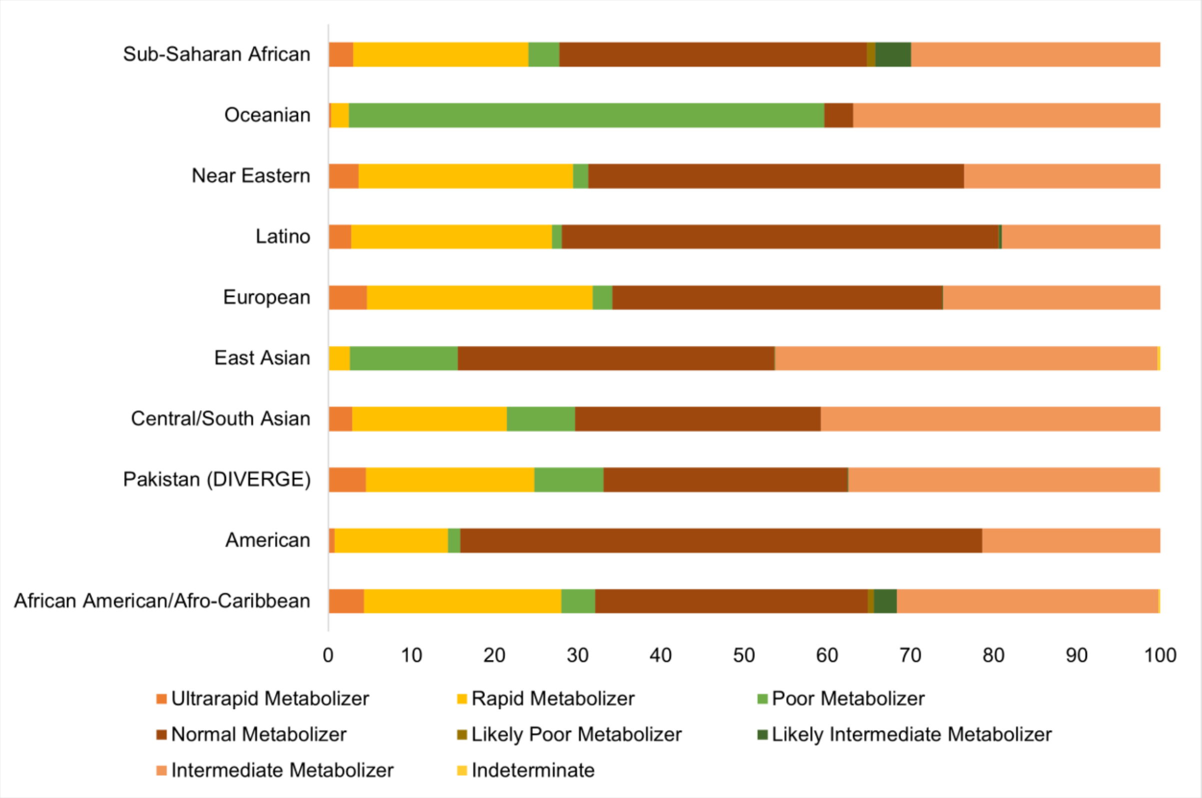

**Supplementary Figure 5:** *CYP2C19* metabolizer phenotype frequency across different ancestral populations. X-axis represents percentage. Colours represent the type of the phenotype. Data for the populations other than Pakistan has been extracted from PharmGKB. DIVERGE is also shown for comparison.

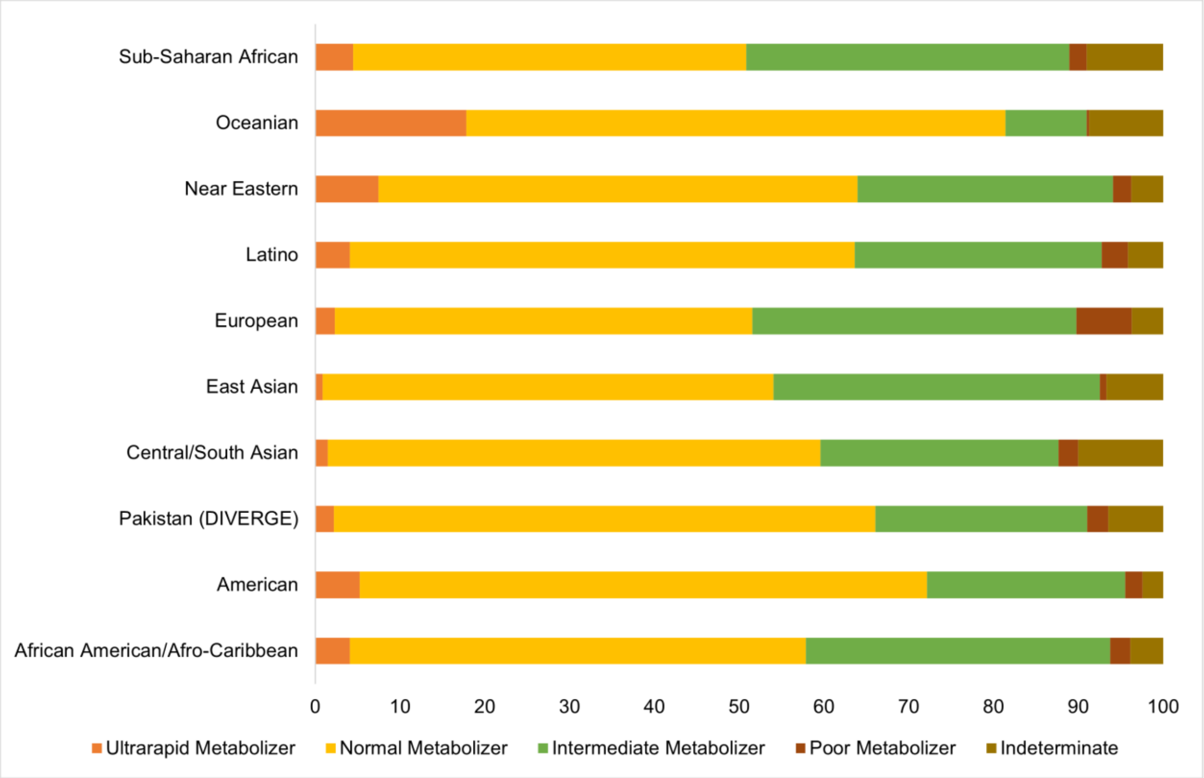

**Supplementary Figure 6:** *CYP2D6* metabolizer phenotype frequency varies across different ancestral populations. X-axis represents percentage. Colours represent the type of the phenotype. Data for the populations other than Pakistan has been extracted from PharmGKB. DIVERGE is also shown for comparison.

#### 4. Supplementary Tables

**Supplementary Table 1: Dichotomizing the response variable**

| **Question:** Overall, since starting treatment for your depression how much have your symptoms changed? | **Status** |
|---|---|
| Complete remission | Non-resistant |
| Partial remission | |
| Episodic symptoms come and go even if I am taking medications | |
| Episodic symptoms relapse after I quit medications | |
| Minimally relieved | Resistant |
| No remission | |

The table shows the question that records response to medications since start of the treatment. This was used to group patients into resistant and non-resistant groups.

**Supplementary Table 2: Selected risk factors for TRD**

| Class | Variable | Instrument/Scale | Type |
|---|---|---|---|
| Demographics | | | |
| | Healthcare setup | Screener | Binary |
| | Age | Screener | Continuous |
| | Gender | Screener | Binary |
| | Provincial ethnicity | Screener | Categorical |
| | Parental relatedness | Screener | Categorical |
| | Marital status | SES | Categorical |
| | Home facilities | SES | Continuous |
| | Education | SES | Binary |
| | Social support | OSSS-3 | Categorical |
| | Life events checklist | LEC-5 | Continuous |
| | Experiences with battering | WEB | Continuous |
| Clinical | | | |
| | Illness duration | DI-PAD | Continuous |
| | Family history of psychiatric disorders | DI-PAD | Binary |
| | Suicidal behavior | SBQ-r | Continuous |
| | Psychotic symptoms | Screener | Binary |
| | Psychiatric comorbidity | Screener | Binary |
| | Physical comorbidity | Screener | Binary |
| Pharmacogenetic* | CYP2C19 phenotype | DRAGEN Array 1.0 | Categorical |
| | CYP2D6 phenotype | DRAGEN Array 1.0 | Categorical |

Table shows different variables in each domain which were selected for investigation as potential risk factors for TRD. It also lists the section of the interview or instrument/Scale used to record/measure the variable. The type of the variable as categorical or continuous is also given. Psychotic symptoms include hallucinations and/or delusions. * Available for a subset of the data.

*Abbreviations: DI-PAD = Diagnostic Interview for Psychosis and Affective Disorders, SBQ-R = Suicidal Behaviour Questionnaire-Revised, LEC-5 = Life Events Checklist-5,SES = Socioeconomic status, WEB= Women Experiences with Battering.*

**Supplementary Table 3:** Demographics of the overall sample and stratification by treatment resistance status

| Variables | Overall (n: 3677) | NTRD (n: 2412) | TRD (n: 1265) | P |
|---|---|---|---|---|
| | **Mean (SD)** | **Mean (SD)** | **Mean (SD)** | |
| Age (Years) | 37·82 (11·07) | 38·33 (11·12) | 36·85 (10·92) | 0·0001 |
| Battering Scale Score | 1·28 (6·53) | 1·39 (6·99) | 1·07 (5·54) | 0·148 |
| Stressful Life Events | 1·70 (2·52) | 1·73 (2·51) | 1·64 (2·56) | 0·303 |
| | **n (%)** | **n (%)** | **n (%)** | |
| Female | 2204 (59·9) | 1460 (60·5) | 744 (58·8) | 0·330 |
| Education Status | 2424 (65·9) | 1584 (65·7) | 840 (66·4) | 0·683 |
| Parental Relatedness | | | | |
| Not Related | 1880 (51·1) | 1182 (49·0) | 698 (55·2) | 0·005 |
| First Cousins | 1370 (37·3) | 935 (38·8) | 435 (34·4) | |
| Second Cousins | 301 (8·2) | 210 (8·7) | 91 (7·2) | |
| Unknown | 126 (3·4) | 85 (3·5) | 41 (3·2) | |
| Provincial Ethnicity (Self-Reported) | | | | |
| Khyber Pakhtunkhwa | 1746 (47·5) | 1108 (45·9) | 638 (50·4) | 0·080 |
| Punjab | 1454 (39·5) | 986 (40·9) | 468 (37·0) | |
| Sindh | 332 (9·0) | 227 (9·4) | 105 (8·3) | |
| Balochistan | 121 (3·3) | 76 (3·2) | 45 (3·6) | |
| Other | 24 (0·7) | 15 (0·6) | 9 (0·7) | |
| Marital Status | | | | |
| Married | 2902 (78·9) | 1935 (80·2) | 967 (76·4) | 0·059 |
| Engaged | 83 (2·3) | 49 (2·0) | 34 (2·7) | |
| Widowed | 156 (4·2) | 102 (4·2) | 54 (4·3) | |
| Divorced/ Separated | 104 (2·8) | 60 (2·5) | 44 (3·5) | |
| Never Married | 432 (11·7) | 266 (11·0) | 166 (13·1) | |
| Social Support | | | | |
| Poor | 1108 (30·1) | 673 (27·9) | 435 (34·4) | 0·0001 |
| Moderate | 1747 (47·5) | 1173 (48·6) | 574 (45·4) | |
| Strong | 822 (22·4) | 566 (23·5) | 256 (20·2) | |

The table provides overview of the sample demographics and non-clinical variables. The continuous variables are reported as mean with standard deviation. The categorical variables are reported as frequency with percentage. The 'Other' category in provincial ethnicity includes Gilgit Baltistan and Kashmiri ethnicity. These were clubbed together because of the small sample size. P corresponds to chi-square or t-test as appropriate.

*Abbreviations: NTRD =Non-treatment resistant depression, TRD = Treatment resistant depression, SD = Standard deviation.*

**Supplementary Table 4:** Clinical characteristics of the overall sample and stratification by treatment resistance status

| Variables | Overall (n: 3677) | NTRD (n: 2412) | TRD (n: 1265) | P |
|---|---|---|---|---|
| | **Mean (SD)** | **Mean (SD)** | **Mean (SD)** | |
| Age at Onset (Years) | 30·52 (10·34) | 30·42 (10·31) | 30·70 (10·41) | 0·435 |
| Duration of Illness (Years) | 7·33 (7·39) | 7·92 (7·70) | 6·20 (6·61) | $1·3 \times 10^{-11}$ |
| Suicidal Behavior Score | 4·16 (3·59) | 4·05 (3·54) | 4·36 (3·66) | 0·013 |
| | **n (%)** | **n (%)** | **n (%)** | |
| Psychiatric Family History | 1423 (38·7) | 1007 (41·7) | 416 (32·9) | $1·9 \times 10^{-06}$ |
| Physical Comorbidity | 1550 (42·2) | 1072 (44·4) | 478 (37·8) | 0·0001 |
| Psychiatric Comorbidity | 1181 (32·1) | 805 (33·4) | 376 (29·7) | 0·027 |
| Psychotic Symptoms | 286 (7·8) | 182 (7·5) | 104 (8·2) | 0·508 |

Table characterizes depression in overall sample and TRD and NTRD groups. Psychiatric comorbidity includes common psychiatric illnesses. Family history was recorded up to 3rd degree for common psychiatric illnesses. Psychotic symptoms include hallucinations and/or delusions P value corresponds to t-test and chi-square test as appropriate.

*Abbreviations: NTRD = Non-treatment resistant depression, TRD = Treatment resistant depression, SD = Standard deviation LEC-5 = Life Events Checklist-5, SBQ-R = Suicidal Behaviour Questionnaire-Revised.*

**Supplementary Table 5a:** Therapies/strategies used for management of depression in overall sample stratified by resistance status

| Drug Class or Combination | Overall (n: 3677) | NTRD (n: 2412) | TRD (n: 1265) | P. |
|---|---|---|---|---|
| | n (%) | n (%) | n (%) | |
| Mood Stabilizer | 380 (10·3) | 252 (10·4) | 128 (10·1) | 0·799 |
| Antipsychotic | 1714 (46·6) | 1148 (47·6) | 566 (44·7) | 0·107 |
| Benzodiazepine | 1587 (43·2) | 1033 (42·8) | 554 (43·8) | 0·598 |
| Anxiolytics | 484 (13·2) | 311 (12·9) | 173 (13·7) | 0·539 |
| Combination Therapy | 1088 (29·6) | 720 (29·9) | 368 (29·1) | 0·659 |
| Augmentation Therapy | 1907 (51·9) | 1276 (52·9) | 631 (49·9) | 0·088 |
| Combination or Augmentation Therapy | 2473 (67·3) | 1636 (67·8) | 837 (66·2) | 0·326 |
| AD With Any Other Psychotropic Drug | 3156 (85·8) | 2061 (85·4) | 1095 (86·6) | 0·384 |
| **Other types of therapies** | | | | |
| Psychotherapy or Counselling | 234 (6·36) | 179 ( 7·4) | 55 ( 4·3) | 0·0003 |
| Visited a faith healer/religious leader | 1790 (48·68) | 1180 (48·9) | 610 (48·2) | 0·712 |
| Traditional medicine or other alternative treatment | 309 (8·4) | 195 ( 8·1) | 114 ( 9·0) | 0·368 |
| Electroconvulsive therapy | 206 (5·6) | 144 ( 6·0) | 62 ( 4·9) | 0·206 |
| Transcranial Magnetic Stimulation (TMS) | 56 (1·52) | 30 ( 1·2) | 26 ( 2·1) | 0·077 |

Table characterizes pharmacological and non-pharmacological treatment received by overall sample stratified by resistant status. Pharmacological therapies received correspond to their medications at the time of interview while the non-pharmacological therapies are reported over the course of whole illness.

*Abbreviations: NTRD = Non-treatment resistant depression, TRD = Treatment resistant depression, Combination therapy = The concomitant use of more than 1 antidepressant. Augmentation therapy = The co-prescription of a mood stabilizer or antipsychotic with antidepressant.*

**Supplementary Table 5b:** Frequency of psychotropic drugs used in medication naïve patient group in DIVERGE

| Drug Class or Combination | Medication Naive (n: 1954) |
|---|---|
| | n (%) |
| Antidepressant | 1922 (98) |
| Mood Stabilizer | 150 (8) |
| Antipsychotic | 696 (36) |
| Benzodiazepine | 1079 (55) |
| Anxiolytics | 207 (11) |
| Combination Therapy | 405 (21)* |
| Augmentation Therapy | 746 (39)* |
| Combination or Augmentation Therapy | 967 (50)* |
| AD With Any Other Psychotropic Drug | 1609 (84)* |

Table reports the drug classes used in medication naïve cohort. These are individuals who received their first prescription for MDD. Some individuals were not prescribed antidepressant on their first visit but other psychotropic medications. *Percentages calculated with respect to the sample which received one antidepressant at least (N=1,922).

*Abbreviations : NTRD = Non-treatment resistant depression, TRD = Treatment resistant depression, Combination therapy = the concomitant use of more than 1 antidepressant. Augmentation therapy = The co-prescription of a mood stabilizer or antipsychotic with antidepressant.*

**Supplementary Table 6:** Frequency and mean dose of individual drugs stratified by treatment resistance status

| Drug Class | Drugs | Frequency (%) | | | Mean Dose (SD) | | |
|---|---|---|---|---|---|---|---|
| | | NTRD (n: 2412) | TRD (n: 1265) | P | NTRD (n: 2412) | TRD (n: 1265) | P |
| Antidepressant | Paroxetine | 378 (15·7) | 210(16·6) | 0·495 | 29·31 (12·17) | 28·37 (10·45) | 0·345 |
| Antidepressant | Clomipramine | 80 (3·3) | 48(3·8) | 0·512 | 70·81 (42·77) | 67·92 (46·76) | 0·721 |
| Antidepressant | Escitalopram | 747 (31) | 407(32·2) | 0·478 | 15·39 (7·91) | 14·97 (7·60) | 0·382 |
| Antidepressant | Fluoxetine | 513 (21·3) | 239(18·9) | 0·098 | 27·87 (11·60) | 28·29 (13·22) | 0·662 |
| Antidepressant | Mirtazapine | 216 (9) | 134(10·6) | 0·122 | 22·94 (10·31) | 22·41 (10·22) | 0·636 |
| Antidepressant | Sertraline | 492 (20·4) | 218(17·2) | 0·023 | 99·51 (54·63) | 86·82 (49·07) | 0·003 |
| Antidepressant | Doxepin | 5 (0·2) | 1(0·1) | 0·628 | 60·00 (37·91) | 50·00 (N/A) | N/A |
| Antidepressant | Dosulepin | 140 (5·8) | 43(3·4) | 0·002 | 83·45 (46·48) | 80·81 (41·49) | 0·739 |
| Antidepressant | Imipramine | 17 (0·7) | 3(0·2) | 0·111 | 67·65 (44·86) | 50·00 (43·30) | 0·536 |
| Antidepressant | Duloxetine | 67 (2·8) | 18(1·4) | 0·013 | 56·57 (35·27) | 54·44 (28·95) | 0·815 |
| Antidepressant | Venlafaxine | 163 (6·8) | 101(8) | 0·193 | 128·91 (60·10) | 120·65 (64·82) | 0·293 |
| Antidepressant | Vortioxetine | 36 (1·5) | 25(2) | 0·340 | 16·11 (6·34) | 13·60 (4·21) | 0·089 |
| Antidepressant | Desvenlafaxine | 23 (1) | 29(2·3) | 0·002 | 106·30 (49·48) | 103·45 (76·68) | 0·878 |
| Antidepressant | Citalopram | 36 (1·5) | 14(1·1) | 0·418 | 17·36 (10·45) | 13·57 (9·08) | 0·239 |
| Antidepressant | Amitriptyline | 110 (4·6) | 65(5·1) | 0·484 | 36·25 (20·87) | 30·06 (15·64) | 0·040 |
| Antidepressant | Trazodone | 24 (1) | 27 (2·1) | 0·008 | 71·88 (55·32) | 75·19 (50·77) | 0·825 |
| Antidepressant | Nortriptyline | 44 (1·8) | 26(2·1) | 0·719 | 31·42 (20·67) | 25·80 (16·29) | 0·247 |
| Antidepressant | Bupropion | 39 (1·6) | 17(1·3) | 0·617 | 188·72 (104·02) | 167·94 (85·44) | 0·473 |
| Antidepressant | Agomelatine | 16 (0·7) | 5(0·4) | 0·427 | 25·94 (6·88) | 25·00 (0·00) | 0·768 |
| Antidepressant | Fluvoxamine | 17 (0·7) | 17 (1·3) | 0·082 | 117·65 (58·47) | 117·65 (49·82) | 1·000 |
| Antidepressant | Moclobemide | 0 (0) | 1 (0·1) | 0·743 | N/A (N/A) | 500·00(N/A) | N/A |

| Drug Class | Drugs | Frequency (%) NTRD (n: 2412) | TRD (n: 1265) | P | Mean Dose (SD) NTRD (n: 2412) | TRD (n: 1265) | P |
|---|---|---|---|---|---|---|---|
| Anxiolytic | Propranolol | 219 (9·1) | 111 (8·8) | 0·805 | 29·90 (20·14) | 30·86(21·27) | 0·689 |
| Anxiolytic | Pregabalin | 83 (3·4) | 52 (4·1) | 0·351 | 100·00 (77·60) | 119·23 (98·22) | 0·209 |
| Anxiolytic | Buspirone | 17 (0·7) | 12 (0·9) | 0·550 | 17·06 (34·54) | 12·08 (3·34) | 0·625 |
| Anxiolytic | Etifoxine | 1 (0) | 3 (0·2) | 0·237 | 100·00 (NA) | 83·33 (28·87) | N/A |
| Antipsychotic | Fluphenazine | 7 (0·3) | 5 (0·4) | 0·821 | 18·79 (17·94) | 6·62 (12·25) | 0·263 |
| Antipsychotic | Quetiapine | 208 (8·6) | 122 (9·6) | 0·333 | 61·66 (54·61) | 76·76 (88·50) | 0·057 |
| Antipsychotic | Olanzapine | 594 (24·6) | 289 (22·8) | 0·246 | 5·56 (3·19) | 5·76 (3·43) | 0·390 |
| Antipsychotic | Paliperidone | 6 (0·2) | 2 (0·2) | 0·851 | 2·25 (0·82) | 3·00 (0·00) | 0·267 |
| Antipsychotic | Risperidone | 199 (8·3) | 90 (7·1) | 0·250 | 1·57 (1·30) | 1·69 (1·16) | 0·424 |
| Antipsychotic | Trifluoperazine | 71 (2·9) | 33 (2·6) | 0·633 | 2·76 (3·25) | 3·71 (5·22) | 0·262 |
| Antipsychotic | Haloperidol | 61 (2·5) | 25 (2) | 0·348 | 4·37 (2·59) | 5·03 (3·76) | 0·355 |
| Antipsychotic | Levosulpiride | 44 (1·8) | 26 (2·1) | 0·719 | 68·75 (27·03) | 74·04 (28·71) | 0·442 |
| Antipsychotic | Clozapine | 4 (0·2) | 3 (0·2) | 0·942 | 62·50 (43·30) | 50·00 (43·30) | 0·721 |
| Antipsychotic | Aripiprazole | 29 (1·2) | 17 (1·3) | 0·833 | 14·93 (7·60) | 13·38 (5·86) | 0·474 |
| Antipsychotic | Lurasidone | 7 (0·3) | 3 (0·2) | 1·000 | 40·00 (16·33) | 33·33 (11·55) | 0·545 |
| Antipsychotic | Chlorpromazine | 1 (0) | 1 (0·1) | 1·000 | 50·00 (N/A) | 100·00 (N/A) | N/A |
| Antipsychotic | Prochlorperazine | 6 (0·2) | 6 (0·5) | 0·404 | 5·83 (2·04) | 6·67 (2·58) | 0·549 |
| Antipsychotic | Ziprasidone | 0 (0) | 1 (0·1) | 0·743 | N/A (N/A) | 80·00 (N/A) | N/A |
| Benzodiazepine | Alprazolam | 319 (13·2) | 148 (11·7) | 0·205 | 0·59 (0·73) | 0·76 (0·98) | 0·036 |
| Benzodiazepine | Clonazepam | 322 (13·3) | 226 (17·9) | <0·001 | 1·12 (0·86) | 1·08 (1·47) | 0·697 |
| Benzodiazepine | Bromazepam | 235 (9·7) | 111 (8·8) | 0·370 | 3·11 (1·02) | 3·20 (1·04) | 0·469 |
| Benzodiazepine | Lorazepam | 34 (1·4) | 9 (0·7) | 0·087 | 3·39 (3·15) | 2·44 (1·01) | 0·383 |
| Benzodiazepine | Diazepam | 147 (6·1) | 66 (5·2) | 0·314 | 5·36 (2·32) | 5·76 (2·23) | 0·244 |

| Drug Class | Drugs | Frequency (%) NTRD (n: 2412) | TRD (n: 1265) | P | Mean Dose (SD) NTRD (n: 2412) | TRD (n: 1265) | P |
|---|---|---|---|---|---|---|---|
| Benzodiazepine | Chlordiazepoxide | 2 (0·1) | 1 (0·1) | 1·000 | 17·50 (10·61) | 10·00 (N/A) | N/A |
| Benzodiazepine | Estazolam | 14 (0·6) | 7 (0·6) | 1·000 | 1·54 (0·57) | 1·86 (0·38) | 0·196 |
| Benzodiazepine | Clobazam | 1 (0) | 0 (0) | 1·000 | 20·00 (N/A) | N/A (N/A) | N/A |
| Mood Stabilizer | Valproic Acid | 85 (3·5) | 48 (3·8) | 0·746 | 564·12 (229·60) | 574·47 (249·54) | 0·810 |
| Mood Stabilizer | Lithium | 33 (1·4) | 11 (0·9) | 0·246 | 512·50 (182·72) | 400·00 (0·00) | 0·049 |
| Mood Stabilizer | Lamotrigine | 91 (3·8) | 41 (3·2) | 0·465 | 127·60 (92·93) | 99·39 (68·80) | 0·084 |
| Mood Stabilizer | Topiramate | 36 (1·5) | 22 (1·7) | 0·667 | 63·19 (41·61) | 62·50 (40·64) | 0·951 |
| Mood Stabilizer | Carbamazepine | 19 (0·8) | 9 (0·7) | 0·958 | 378·95 (257·29) | 400·00 (244·95) | 0·839 |
| Mood Stabilizer | Gabapentin | 9 (0·4) | 5 (0·4) | 1·000 | 241·67 (365·72) | 170·00 (97·47) | 0·680 |
| Sedative | Zolpidem | 9 (0·4) | 10 (0·8) | 0·151 | 9·17 (2·50) | 8·75 (2·70) | 0·732 |
| Sedative | Eszopiclone | 8 (0·3) | 5 (0·4) | 0·987 | 2·00 (0·76) | 1·80 (0·45) | 0·606 |

Table reports on the classification of individual drugs in different drug classes. It also shows the frequency of each drug and its average dose stratified by TRD status to provide a comparison. P compares frequency and mean doses of drugs between TRD and NTRD using chi-square and t-test respectively.

*Abbreviations: NTRD = Non-treatment resistant depression, TRD = Treatment resistant depression.*

**Supplementary Table 7: Parameters showing model performance**

| Parameter | Type | ICC | AIC | BIC | R2 |
|---|---|---|---|---|---|
| M0_glm | Null simple regression | - | 4736 | 4742 | - |
| M0 | Null ML regression | 0·79 | 4459 | 4472 | - |
| M1_demog | ML regression with demographic predictors | - | 4386 | 4528 | 0·14 |
| M1_clin | ML regression with clinical predictors | - | 4412 | 4524 | 0·11 |
| M2 | ML regression with both clinical and demographic predictors | - | 4381·8 | 4555·7 | 0·14 |
| M2_pgx* | M2 plus CYP enzyme phenotype predictors | - | 1303 | - | 0·17 |

Table reports model performance parameters for different models. *Effective sample size for this analysis is 1,085. For the rest its 3,677.

*Abbreviations: M0_glm = simple regression model with no predictors, M0 = Null multilevel model with random effects added for Site. M1_demog = Multivariate model with level demographic predictors, M1_clin = Multivariate model with level demographic predictors. M2 = Demographic and clinical predictors, R2 = R-squared, ICC = Intra-class correlation coefficient.*

**Supplementary Table 8:** Predictors of social support

| Variables | OR (95% CI) | P |
|---|---|---|
| Partners Routinely Separated | 0·99 (0·84 - 1·18) | 0·936 |
| Relative Live in Same Area | 4·20 (3·60 - 4·95) | $3{\cdot}9 \times 10^{-71}$ |
| Family Structure (Binary Family) | Ref· | |
| Living Alone | 0·74 (0·36 - 1·48) | 0·392 |
| Extended Family (In-Laws) | 1·22 (1·05 - 1·43) | 0·012 |
| Extended Family (Parents) | 1·32 (1·13 - 1·55) | $4{\cdot}5 \times 10^{-4}$ |
| Extended Family (GPs, Uncles w/o Partial Separation) | 1·98 (1·42 - 2·77) | $6{\cdot}2 \times 10^{-5}$ |
| Extended Family (GPs, Uncles with Partial Separation) | 1·29 (0·96 - 1·73) | 0·088 |

Table shows results from association analysis between predictors of social support and social support (OSSS-3) as outcome. Level of significance is set to p<0·01 (Bonferroni adjustment).

*Abbreviations: GPs = Grandparents.*

**Supplementary Table 9a*:*** Demographic and clinical risk factors for TRD in a combined model

| Variables | UOR (95% CI) | P | AOR (95% CI) | P |
|---|---|---|---|---|
| Age (Years) | 0·98 (0·98-0·99) | $1{\cdot}2\text{x}10^{-5}$ | 1·00 (0·99-1·00) | 0·344 |
| Duration Of Illness (Years) | 0·96 (0·95-0·98) | $1{\cdot}9\text{x}10^{-10}$ | 0·97 (0·95-0·98) | $4{\cdot}4 \text{ x } 10^{-8}$ |
| Female Gender | 0·90 (0·77–1·05) | 0·179 | 0·88 (0·74-1·04) | 0·134 |
| Setup Type | - | - | 0·90 (0·56-1·47) | 0·686 |
| Psychiatric Family History | 0·82 (0·70-0·96) | 0·015 | 0·83 (0·71-0·98) | 0·027 |
| Physical Comorbidity | 0·91 (0·78-1·06) | 0·229 | 0·99 (0·84-1·18) | 0·947 |
| Psychiatric Comorbidity | 1·06 (0·90-1·25) | 0·457 | 1·05 (0·88-1·24) | 0·595 |
| Psychotic Symptoms | 1·41 (1·07-1·85) | 0·015 | 1·35 (1·01-1·79) | 0·043 |
| Educated | 1·03 (0·88-1·21) | 0·690 | 0·98 (0·82-1·18) | 0·829 |
| Home Facilities Score | 0·94 (0·90-0·99) | 0·016 | 0·96 (0·92-1·02) | 0·176 |
| Battering Score | 1·00 (0·99-1·01) | 0·962 | 1·00 (0·98-1·01) | 0·594 |
| Stressful Life Events Score | 1·03 (1·00-1·06) | 0·053 | 1·02 (0·99-1·06) | 0·131 |
| Suicidal Behavior | 1·03 (1·01-1·05) | 0·002 | 1·02 (1·00-1·05) | 0·029 |
| Social Support (Poor) | Ref· | | Ref· | |
| Moderate | 0·64 (0·53-0·76) | $5{\cdot}7\text{x}10^{-7}$ | 0·64 (0·54-0·77) | $2{\cdot}2\text{x}10^{-6}$ |
| Strong | 0·54 (0·44-0·68) | $4{\cdot}5\text{x}10^{-8}$ | 0·56 (0·45-0·70) | $4{\cdot}2\text{x}10^{-7}$ |
| Parental Relatedness (Unrelated) | Ref· | | Ref· | 0·12 * |
| First Cousins | 0·84 (0·71-0·99) | 0·036 | 0·81 (0·69-0·96) | 0·016 |
| Second Cousins | 0·87 (0·65-1·15) | 0·337 | 0·85 (0·63-1·13) | 0·262 |
| Unknown | 0·87 (0·57-1·29) | 0·485 | 0·87 (0·57-1·32) | 0.512 |
| Provincial Ethnicity (Khyber Pakhtunkhwa) | Ref. | | Ref. | 0.39 * |
| Punjab | 0·82 (0·57-1·17) | 0·267 | 0·85 (0·59-1·24) | 0·404 |
| Sindh | 0·78 (0·46-1·30) | 0·345 | 0·74 (0·43-1·26) | 0·271 |
| Baluchistan | 1·49 (0·82-2·73) | 0·189 | 1·37 (0·74-2·54) | 0·31 |
| Other | 1·38 (0·54-3·35) | 0·486 | 1·45 (0·57-3·67) | 0.437 |
| Marital Status (Married) | Ref. | | Ref. | 0·05 * |
| Engaged | 1·59 (0·99-2·52) | 0·051 | 1·30 (0·80-2·12) | 0·289 |
| Widowed | 1·10 (0·76-1·57) | 0·600 | 1·30 (0·89-1·89) | 0·177 |
| Divorced/Separated | 1·80 (1·18-2·72) | 0·006 | 1·81 (1·18-2·79) | 0·007 |
| Never Married | 1·32 (1·06-1·64) | 0·014 | 1·14 (0·89-1·46) | 0·305 |

Table reports regression results for different risk factors for TRD as outcome (N = 3,677). Setup Type = Private or public sector recruitment centre P corresponds to the multilevel logistic regression. *Indicates p value from the LRT test comparing different models. A basic model (supplementary table 8b) containing all other variables except parental relatedness, provincial ethnicity and marital status was compared against basic model plus each of these three variables separately to improve the goodness of fit. Psychotic symptoms refer to either hallucinations or delusions or both.

*Abbreviations: OR = Odds ratio, 95% CI = 95% Confidence interval, UOR = Unadjusted odds ratio, AOR = adjusted odds ratio*

**Supplementary Table 9b:** Basic model of TRD risk factors

| Variables | UOR (95% CI) | P | AOR (95% CI) | P |
|---|---|---|---|---|
| Age (Years) | 0·98 (0·99-0·98) | $1{\cdot}2 \times 10^{-5}$ | 0·99 (0·99-1·00) | 0·173 |
| Duration of Illness (Years) | 0·96 (0·98-0·95) | $1{\cdot}9 \times 10^{-10}$ | 0·97 (0·96-0·98) | $1 \times 10^{-7}$ |
| Female Gender | 0·90 (1·05-0·77) | 0·179 | 0·89 (0·75-1·05) | 0·177 |
| Setup Type | | | 0·90 (0·56-1·46) | 0·677 |
| Psychiatric Family History | 0·82 (0·96-0·70) | 0·015 | 0·83 (0·71-0·98) | 0·027 |
| Physical Comorbidity | 0·91 (1·06-0·78) | 0·229 | 0·99 (0·83-1·17) | 0·868 |
| Psychiatric Comorbidity | 1·06 (1·25-0·90) | 0·457 | 1·05 (0·89-1·25) | 0·569 |
| Psychotic Symptoms (Hallucinations or Delusions) | 1·41 (1·85-1·07) | 0·015 | 1·34 (1·01-1·78) | 0·046 |
| Suicidal Behavior Score | 1·03 (1·05-1·01) | 0·002 | 1·02 (1·00-1·05) | 0·029 |
| Education Status | 1·03 (1·21-0·88) | 0·690 | 0·99 (0·83-1·19) | 0·956 |
| Home Facilities Score | 0·94 (0·99-0·90) | 0·016 | 0·97 (0·92-1·02) | 0·209 |
| Battering Score | 1·00 (1·01-0·99) | 0·962 | 1·00 (0·98-1·01) | 0·542 |
| Stressful Life Events Score | 1·03 (1·06–1·00) | 0·053 | 1·03 (1·00-1·06) | 0·059 |
| Social Support (Poor) | Ref. | | Ref. | |
| Moderate | 0·64 (0·76-0·53) | $5{\cdot}7 \times 10^{-7}$ | 0·64 (0·53-0·77) | $1{\cdot}2 \times 10^{-6}$ |
| Strong | 0·54 (0·68-0·44) | $4{\cdot}5 \times 10^{-8}$ | 0·55 (0·44-0·69) | $1{\cdot}2 \times 10^{-7}$ |

Table shows a multivariate model for risk factors of TRD. Regression results for different risk factors for TRD as outcome are shown (N = 3,677). This model excludes provincial ethnicity, marital status and parental relatedness.

*Abbreviations: OR= Odds ratio, 95% CI = 95% Confidence interval, UOR = Unadjusted odds ratio, AOR = adjusted odds ratio. P corresponds to the multilevel logistic regression.*

**Supplementary Table 10:** Overall metabolizer frequency for CYP2C19 and CYP2D6 in Pakistan

| Metabolizer Status | CYP2C19 (N=3,217) Frequency (%) | CYP2D6 (N=3,154) Frequency (%) |
|---|---|---|
| Intermediate Metabolizer | 1204 (37·43) | 789 (25·02) |
| Normal Metabolizer | 942 (29·28) | 2013 (63·82) |
| Rapid Metabolizer | 651 (20·24) | -- |
| Poor Metabolizer | 268 (8·33) | 78 (2·47) |
| Ultrarapid Metabolizer | 146 (4·54) | 70 (2·22) |
| Likely Intermediate Metabolizer | 4 (0·12) | -- |
| Indeterminate | 2 (0·06) | 204 (6·47) |
| No Call | -- | 63 (2·00) |

Table shows the prevalence of metabolic phenotype for the two selected genes. For *CYP2D6* the effective sample size is less than 3,217 because 2% of the subjects had failed calls for metaboliser status for *CYP2D6*.

**Supplementary Table 11:** Star allele frequency for *CYP2C19* in the Pakistani population

| Star Allele | % Frequency (N=3,217) |
|---|---|
| *1 | 41·98 |
| *2 | 26·66 |
| *17 | 19·51 |
| *38 | 11·33 |
| *3 | 0·28 |
| *8 | 0·06 |
| *39 | 0·03 |
| *4 | 0·03 |
| *10 | 0·02 |
| *19 | 0·02 |
| *24 | 0·02 |
| *25 | 0·02 |
| *26 | 0·02 |
| *32 | 0·02 |
| *35 | 0·02 |
| *9 | 0·02 |

**Supplementary Table 12:** Star allele frequency for *CYP2D6* in the Pakistani population

| Star Allele | % Frequency (N=3,153) | Star Allele | % Frequency (N=3,153) | Star Allele | % Frequency (N=3,153) |
|---|---|---|---|---|---|
| *1 | 38·95 | *24+*13 | 0·11 | *75 | 0·03 |
| *2 | 32·99 | *65 | 0·11 | *8 | 0·03 |
| *4 | 8·05 | *160 | 0·08 | *89 | 0·03 |
| *10 | 3·15 | *34 | 0·08 | *10+*68 | 0·02 |
| *5 | 2·46 | *4N+*4 | 0·08 | *1+*121 | 0·02 |
| *68+*4 | 2·44 | *74 | 0·08 | *1+*24 | 0·02 |
| *41 | 1·52 | *1+*68 | 0·06 | *1+*2x2 | 0·02 |
| *24 | 0·94 | *22 | 0·06 | *134 | 0·02 |
| *86 | 0·86 | *36 | 0·06 | *1+*36 | 0·02 |
| *33 | 0·84 | *116 | 0·05 | *1+*36x3 | 0·02 |
| *35 | 0·68 | *121 | 0·05 | *150 | 0·02 |
| *1x2 | 0·67 | *1+*68+*4 | 0·05 | *17+*13 | 0·02 |
| *7 | 0·65 | *36x2+*10 | 0·05 | *19 | 0·02 |
| *2x2 | 0·6 | *4+*13 | 0·05 | *1x3+*36 | 0·02 |
| *111 | 0·54 | *108 | 0·03 | *1x5 | 0·02 |
| *13 | 0·33 | *10x2 | 0·03 | *20 | 0·02 |
| *13+*1 | 0·29 | *117 | 0·03 | *24+*68+*4 | 0·02 |
| *36+*10 | 0·27 | *131 | 0·03 | *29 | 0·02 |
| *13+*2 | 0·25 | *132 | 0·03 | *53 | 0·02 |
| *6 | 0·25 | *143 | 0·03 | *59 | 0·02 |
| *68 | 0·22 | *146 | 0·03 | *65+*36+*10 | 0·02 |
| *9 | 0·22 | *2+*65 | 0·03 | *68+*4x2 | 0·02 |
| *112 | 0·17 | *2+*68 | 0·03 | *7+*13 | 0·02 |
| *3 | 0·17 | *28 | 0·03 | *7+*130 | 0·02 |
| *4x2 | 0·16 | *33x2 | 0·03 | *7+*149 | 0·02 |
| *17 | 0·14 | *40 | 0·03 | *73 | 0·02 |
| *43 | 0·14 | *68x2 | 0·03 | *88 | 0·02 |
| *39 | 0·13 | *6x2 | 0·03 | | |

**Supplementary Table 13:** Provincial prevalence of *CYP2C19* metaboliser phenotype

| Phenotype | Provincial Ethnicity | Provincial Frequency | Provincial Prevalence |
|---|---|---|---|
| Ultrarapid Metabolizer | Khyber Pakhtunkhwa | 67 | 5·64 |
| | Punjab | 74 | 4·36 |
| | Sindh | 4 | 1·47 |
| | Balochistan | - | - |
| | Other | 1 | 8·33 |
| Rapid Metabolizer | Khyber Pakhtunkhwa | 272 | 22·91 |
| | Punjab | 313 | 18·46 |
| | Sindh | 52 | 19·12 |
| | Balochistan | 11 | 22·00 |
| | Other | 3 | 25·00 |
| Poor Metabolizer | Khyber Pakhtunkhwa | 66 | 5·56 |
| | Punjab | 178 | 10·50 |
| | Sindh | 23 | 8·46 |
| | Balochistan | 1 | 2·00 |
| Normal Metabolizer | Khyber Pakhtunkhwa | 385 | 32·43 |
| | Punjab | 458 | 27·00 |
| | Sindh | 83 | 30·51 |
| | Balochistan | 14 | 28·00 |
| | Other | 2 | 16·67 |
| Likely Intermediate Metabolizer | Khyber Pakhtunkhwa | 1 | 0·08 |
| | Punjab | 1 | 0·06 |
| | Sindh | 1 | 0·37 |
| | Balochistan | 1 | 2·00 |
| Intermediate Metabolizer | Khyber Pakhtunkhwa | 396 | 33·36 |
| | Punjab | 670 | 39·50 |
| | Sindh | 109 | 40·07 |
| | Balochistan | 23 | 46·00 |
| | Other | 6 | 50·00 |
| Indeterminate | Punjab | 2 | 0·12 |

Participants per each provincial group: Khyber Pakhtunkhwa = 1187, Punjab = 1696, Sindh = 272, Balochistan = 50, Other = 12.

**Supplementary Table 14:** Provincial prevalence of *CYP2D6* metaboliser phenotype

| Phenotype | Provincial Ethnicity | Frequency | Prevalence |
|---|---|---|---|
| Ultrarapid Metabolizer | Khyber Pakhtunkhwa | 22 | 1·89 |
| | Punjab | 39 | 2·35 |
| | Sindh | 7 | 2·59 |
| | Balochistan | 1 | 2·00 |
| | Other | 1 | 8·33 |
| Poor Metabolizer | Khyber Pakhtunkhwa | 28 | 2·40 |
| | Punjab | 37 | 2·23 |
| | Sindh | 13 | 4·81 |
| Normal Metabolizer | Khyber Pakhtunkhwa | 742 | 63·69 |
| | Punjab | 1051 | 63·43 |
| | Sindh | 176 | 65·19 |
| | Balochistan | 36 | 72·00 |
| | Other | 8 | 66·67 |
| Intermediate Metabolizer | Khyber Pakhtunkhwa | 300 | 25·75 |
| | Punjab | 411 | 24·80 |
| | Sindh | 66 | 24·44 |
| | Balochistan | 11 | 22·00 |
| | Other | 1 | 8·33 |
| Indeterminate | Khyber Pakhtunkhwa | 73 | 6·27 |
| | Punjab | 119 | 7·18 |
| | Sindh | 8 | 2·96d |
| | Balochistan | 2 | 4·00 |
| | Other | 2 | 16·67 |

Participants per each provincial group: Khyber Pakhtunkhwa = 1165, Punjab = 1657, Sindh = 270, Balochistan = 50, Other = 12.

**Supplementary Table 15:** Association of number of reported side effects with *CYP2C19* and *CYP2D6* phenotype

| Variables | LogOdds | Std. Error | P |
|---|---|---|---|
| CYP2C19 Phenotype (Normal Metabolizer) | Ref. | - | - |
| Poor Metabolizer | -0·022 | 0·133 | 0·869 |
| Intermediate Metabolizer | 0·013 | 0·085 | 0·877 |
| Rapid Metabolizer | -0·141 | 0·101 | 0·164 |
| Ultra rapid Metabolizer | 0·231 | 0·168 | 0·169 |
| CYP26 Phenotype (Normal Metabolizer) | Ref. | | |
| Poor Metabolizer | -0·667 | 0·375 | 0·075 |
| Intermediate Metabolizer | 0·062 | 0·123 | 0·611 |
| Ultra rapid | -0·516 | 0·420 | 0·219 |

Table shows results of association analysis between CYP enzymes phenotype and number of sideeffects. Outcome was the number of side-effects to the current medications. This was tested using individuals from TRD dataset who had metaboliser phenotypes available. P value corresponds to the negative binomial regression model.
